# Sperm sequencing reveals extensive positive selection in the male germline

**DOI:** 10.1101/2024.10.30.24316414

**Authors:** Matthew DC Neville, Andrew RJ Lawson, Rashesh Sanghvi, Federico Abascal, My H Pham, Alex Cagan, Pantelis A Nicola, Tetyana Bayzetinova, Adrian Baez-Ortega, Kirsty Roberts, Stefanie V. Lensing, Sara Widaa, Raul E Alcantara, María Paz García, Sam Wadge, Michael R Stratton, Peter J Campbell, Kerrin Small, Iñigo Martincorena, Matthew E Hurles, Raheleh Rahbari

**Affiliations:** Cancer, Ageing and Somatic Mutation, Wellcome Sanger Institute, Hinxton, United Kingdom; Sequencing Operations, Wellcome Sanger Institute, Hinxton, United Kingdom; Quotient Therapeutics Limited, Saffron Walden, UK; Kings College London, Department of Twin Research & Genetic Epidemiology, London, United Kingdom

## Abstract

Mutations that occur in the cell lineages of sperm or eggs can be transmitted to offspring. In humans, positive selection of driver mutations during spermatogenesis is known to increase the birth prevalence of certain developmental disorders. Until recently, characterising the extent of this selection in sperm has been limited by the error rates of sequencing technologies. Using the duplex sequencing method NanoSeq, we sequenced 81 bulk sperm samples from individuals aged 24 to 75 years. Our findings revealed a linear accumulation of 1.67 (95% CI = 1.41-1.92) mutations per year per haploid genome, driven by two mutational signatures associated with human ageing. Deep targeted and exome NanoSeq of sperm samples identified over 35,000 germline coding mutations. We detected 40 genes (31 novel) under significant positive selection in the male germline, implicating both activating and loss-of-function mechanisms and diverse cellular pathways. Most positively selected genes are associated with developmental or cancer predisposition disorders in children, while four genes that exhibit elevated frequencies of protein-truncating variants in healthy populations. We find that positive selection during spermatogenesis drives a 2-3 fold elevated risk of known disease-causing mutations in sperm, resulting in 3-5% of sperm from middle-aged to elderly individuals carrying a pathogenic mutation across the exome. These findings shed light on the dynamics of germline mutations and highlight a broader increased disease risk for children born to fathers of advanced age than previously appreciated.

## Introduction

Human cells in all tissues accumulate mutations throughout life. In replicating tissues, acquired driver mutations that confer a selective advantage can promote the expansion of individual clones within competing stem and progenitor cell populations. While patterns of selection and clonal expansion have been extensively studied in cancers, recent research has also highlighted their occurrence in normal tissues during ageing^1–10^.

The spermatogonial stem cells of the testis occupy a unique niche amongst other studied normal tissues. Among replicating cells, they have the lowest mutation rate, ∼5-20 fold lower than any other studied somatic cell type^7^. They are also the only replicating cells with the potential to transmit mutations to offspring, balancing self-renewal and spermatogenesis to produce 150-275 million sperm per day post-puberty^11,12^. Targeted sequencing studies have revealed that driver mutations are acquired in spermatogonial stem cells and that these cell populations expand along seminiferous tubules, resulting in elevated fractions of mutant clones that are detectable in sperm^13–17^. Interestingly, all germline driver mutations identified so far are activating missense hotspot mutations, which contrasts with a broader range of activating and inactivating driver mutations observed in cancers and somatic tissues. These germline driver mutations can have profound implications for offspring, as they are found in a set of 13 genes all known to cause severe developmental disorders^18^. This leads to a significant increase, up to 1,000-fold, in the sporadic birth prevalence of these disorders, with a strong correlation to elevated age of the father^19^.

Technical limitations, related to the polyclonality and low mutation rate of testis and sperm, have prevented extensive characterisation of this selection beyond a limited set of genes^18^. However, recent advances in error-corrected duplex DNA sequencing approaches, in which information from both DNA strands is used to detect mutations at single molecule resolution^20–22^, have proven successful for the accurate estimation of mutation burden in sperm^23–25^. Here we combine the duplex approaches of whole genome NanoSeq^23^ with deep whole exome and targeted NanoSeq (Lawson A.R., Abascal F., P.A. Nicola et al., manuscript submitted for publication) to characterise positive selection in the male germline and quantify its consequences for accumulation of disease mutations in sperm.

## Results

### Cohort and sequencing coverage

We performed restriction enzyme based, whole genome NanoSeq^23^ of bulk semen samples (n = 81; 1-2 timepoints per donor; age range: 24-75 years) and matched blood (n = 119; 1-3 timepoints; age range: 22-83 years) from 63 men in the TwinsUK cohort^26^ (including 9 monozygotic and 3 dizygotic twin pairs; **Methods**; **Supplementary Table 1**). The analysed sperm samples had sperm counts above 1 million/mL, as those below this threshold showed evidence of somatic cell contamination (**Extended Data Fig. 1**; **Supplementary Note 1**). Across these samples, the mean number of unique DNA molecules per site where a mutation was callable (duplex coverage - dx) was 3.7dx in sperm, and 4.3dx in blood (**Extended Data Fig. 2a**). For sperm, a haploid cell, 1dx is equivalent to one cell, whereas for blood, a diploid cell, 2dx is equivalent to one cell.

### Mutational burden and signatures

We performed stringent variant filtering of single nucleotide variants (SNVs) and small insertion– deletion mutations (indels) from sperm and blood whole genome NanoSeq (**Methods**). From the 6,653 SNVs detected in sperm, we estimated an age-related accumulation of 1.67 substitutions per year per haploid genome (95% CI 1.41-1.92, linear mixed-effect regression). This is comparable to estimates from paternal *de novo* mutations (DNMs) in family pedigrees^27^ of 1.44 substitutions per year (95% CI 1.00-1.87) and seminiferous tubules of the testes^7^ of 1.40 substitutions per year (95% CI 1.02-1.76; **Fig. 1a**). Indels accumulated in sperm at a rate of 0.10 indels per year per haploid genome (95% CI 0.06- 0.15), similar to the rate observed in paternally phased DNMs^27^ of 0.08 haploid indels per year (95% CI -0.02-0.17) and seminiferous tubules of the testes^7^ of 0.08 haploid indels per year (95% CI 0.02- 0.13; **Fig. 1b**).

**Figure 1.**
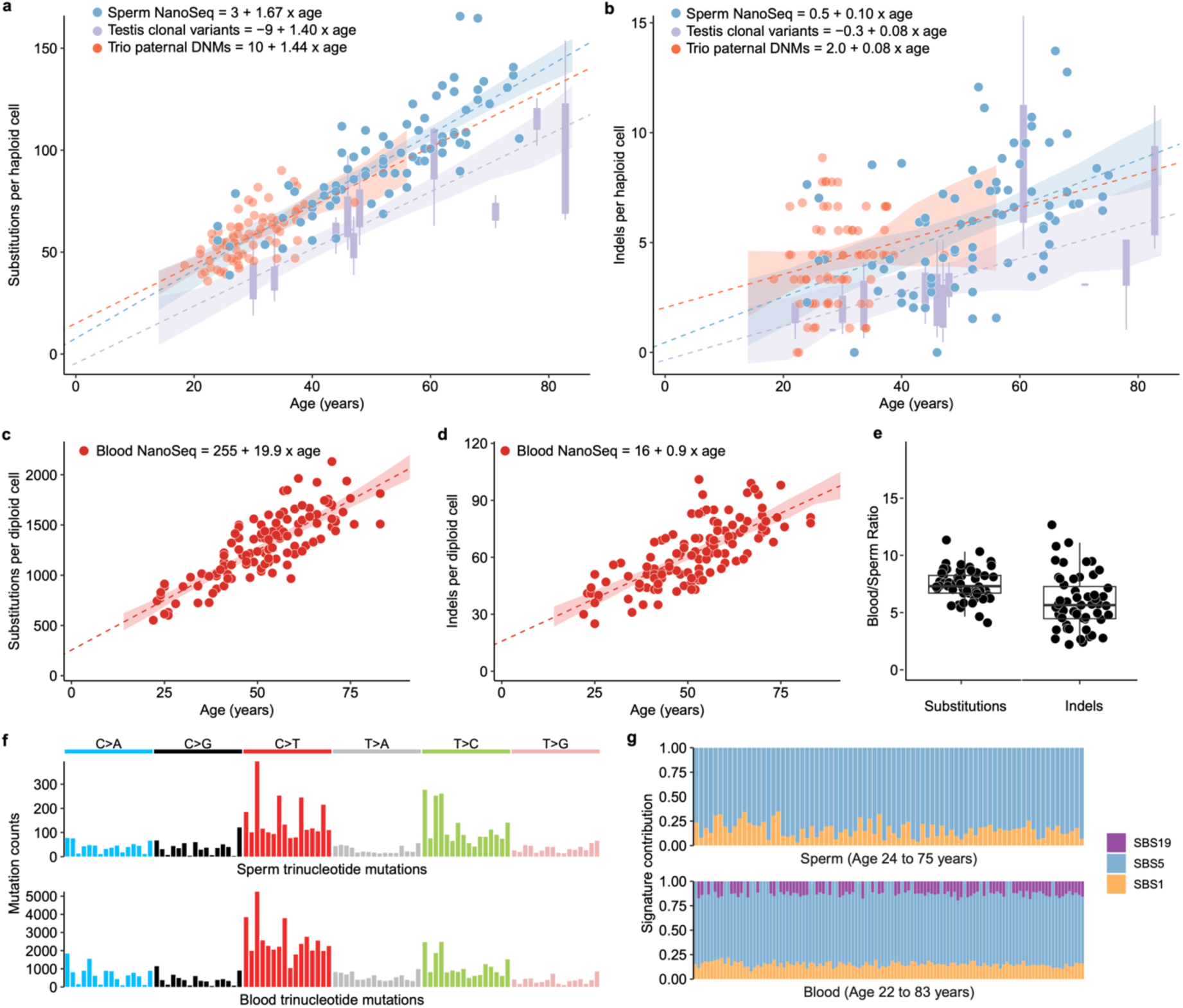
Mutational burden and signature analysis in sperm and matched blood. **a,b,** Substitutions (**a**) and indels (**b**) per haploid cell from sperm whole genome NanoSeq, trio paternal DNMs^27^ called with standard sequencing and clonal variants from seminiferous tubules of testis^7^ called with standard sequencing. Dots indicate single donors while boxplots for testis variants show 1-15 samples per donor. **c,d,** Substitutions (**c**) and indels (**d**) per diploid cell for different ages from blood NanoSeq samples. **e,** Ratio blood to sperm substitutions and indels per diploid cell per year. Each dot corresponds to an individual with both a blood and sperm sample and where individuals had multiple timepoints the mean value of all timepoints in that tissue was used. **f,** Trinucleotide mutation counts in all sperm and blood samples. **g**, Contribution of signatures SBS1, SBS5, and SBS19 in sperm and blood samples ordered by age. **a,b,c,d,** Models are linear mixed regressions with 95% CIs calculated by parametric bootstrapping. **a,b,e,** Box plots show the interquartile range, median, and 95% confidence interval for the median.

From the 92,035 SNVs and 4,641 indels detected in whole blood, we estimated an age-related accumulation of 19.9 substitutions per year per diploid genome (95% CI 17.3-22.5; **Fig. 1c**) and 0.9 indels per year (95% CI 0.7-1.1; **Fig. 1d**). Both estimates are within the range of mutation rates observed for specific cell types in the blood^8^, consistent with measuring a weighted average of these cell types in whole blood (**Extended Data Fig. 3a,b**). We find that individuals had a mean of 7.6-fold more substitutions per bp per year (range 4.2-11.5; **Fig. 1e**) and 6.3-fold more indels per bp per year (range 2.2-18.7; **Fig. 1e**) in blood than in sperm. Accounting for twin status or multiple timepoints from the same individuals had a significant predictive effect for mutation burden in blood but not in sperm (**Supplementary Note 2**).

The SNV mutational signatures in sperm were inferred to be SBS1 (mean 16%) and SBS5 (mean 84%), the expected clock-like ageing signatures^28^ (**Fig. 1f-g**). In blood, SBS1 (mean 15%) and SBS5 (mean 75%) were also the main mutational signatures, with an additional contribution of SBS19 (mean 10%), which has been linked to persistent DNA lesions in hematopoietic stem cells^28^ (**Fig. 1f-g)**. We observed that all signatures were correlated with age (**Extended Data Fig. 3c,d**). SBS1 and SBS5 accumulated in individuals at a mean of 8.9-fold (range 2.3-39.1) and 6.8-fold (range 3.7-10.9) higher rate in blood than in sperm respectively (**Extended Data Fig. 3e**), indicating that SBS19 does not explain a substantial fraction of the mutation burden gap between the two tissues.

### Selective pressure dynamics in sperm

To investigate positive selection in protein-coding regions in sperm we required much greater duplex coverage. Therefore, we utilised a capture-based modification to NanoSeq (Lawson A.R., Abascal F., P.A. Nicola et al., manuscript submitted for publication) to deeply sequence coding regions from the same set of semen samples. Specifically, we sequenced 38 samples using whole-exome NanoSeq to a mean depth of 551dx per sample (20,923 cumulative dx), and 81 samples using targeted NanoSeq to a mean depth of 985dx per sample (79,811 cumulative dx) with a target panel consisting of 263 canonical cancer driver genes, 107 of which are also associated with developmental disorders (**Extended Data Fig. 2a; Supplementary Table 2; Methods**). After variant filtering **(Methods**), we detected 56,503 (58% within coding regions) SNV/indel mutations from the exome panel and 5,059 (58% within coding regions) from the targeted cancer panel. The age correlation of mutation burden for exome and targeted sample sets were consistent with whole genome NanoSeq after correcting for the relative trinucleotide composition of sequencing coverage (**Extended Data Fig. 2b**).

The vast majority of variants (99.5%) were detected only in a single duplex molecule of a sample. Similarly, in the 23 samples with two timepoints (mean 12.1 year gap), 99.3% of the 5,143 variant calls from the first timepoint were not called in the second timepoint. These results are consistent with sperm being a highly polyclonal collection of cells derived from a large population of spermatogonial stem cell progenitors in the testis.

The exome-wide strength of positive selection in sperm was quantified by estimating the rate of non-synonymous (*N*) relative to selectively neutral synonymous (*S*) mutations (dN/dS ratio, where dN/dS = 1.0 indicates neutrality). We employed the *dNdScv* algorithm, which by default calculates dN/dS while adjusting for trinucleotide context and several gene-level genomic covariates that influence mutation rate^29^. We modified this algorithm in three ways: first, we adjusted for duplex sequencing coverage per base to correct for differential coverage within and between genes; second, we incorporated an adjustment for CpG methylation levels in the testis due to its significant influence on mutation rates; and third, we switched from trinucleotide to pentanucleotide context to better account for the effects of extended contexts on germline mutation rates^30^. These modifications refined exome-wide dN/dS ratios by resolving specific mutation rate biases but had minor effects on gene-level dN/dS ratios (**Extended Data Fig. 4, Supplementary Note 3**).

Using this model, we estimated the dN/dS ratio in the exome-sequenced samples to be 1.07 (95% CI 1.04-1.10). This ratio implies that 6.5% (95% CI 3.8%-9.1%) of the observed non-synonymous substitutions in sperm conferred a clonal advantage during spermatogenesis in this cohort. Splitting the cohort into thirds by age, we find that the exome-wide dN/dS ratio increased with age. The ratio was 1.01 (95% CI 0.93-1.09) in 26-42 year olds, 1.03 (95% CI 0.97-1.10) in 43-58 year olds, and 1.09 (95% CI 1.06-1.13) in 59-74 year olds (**Fig. 2f**). This suggests that the dN/dS ratio increases over male lifespan and that the cohort wide dN/dS ratios presented here in part reflect the age distribution of samples (age range 26-74; mean 53 years).

**Figure 2.**
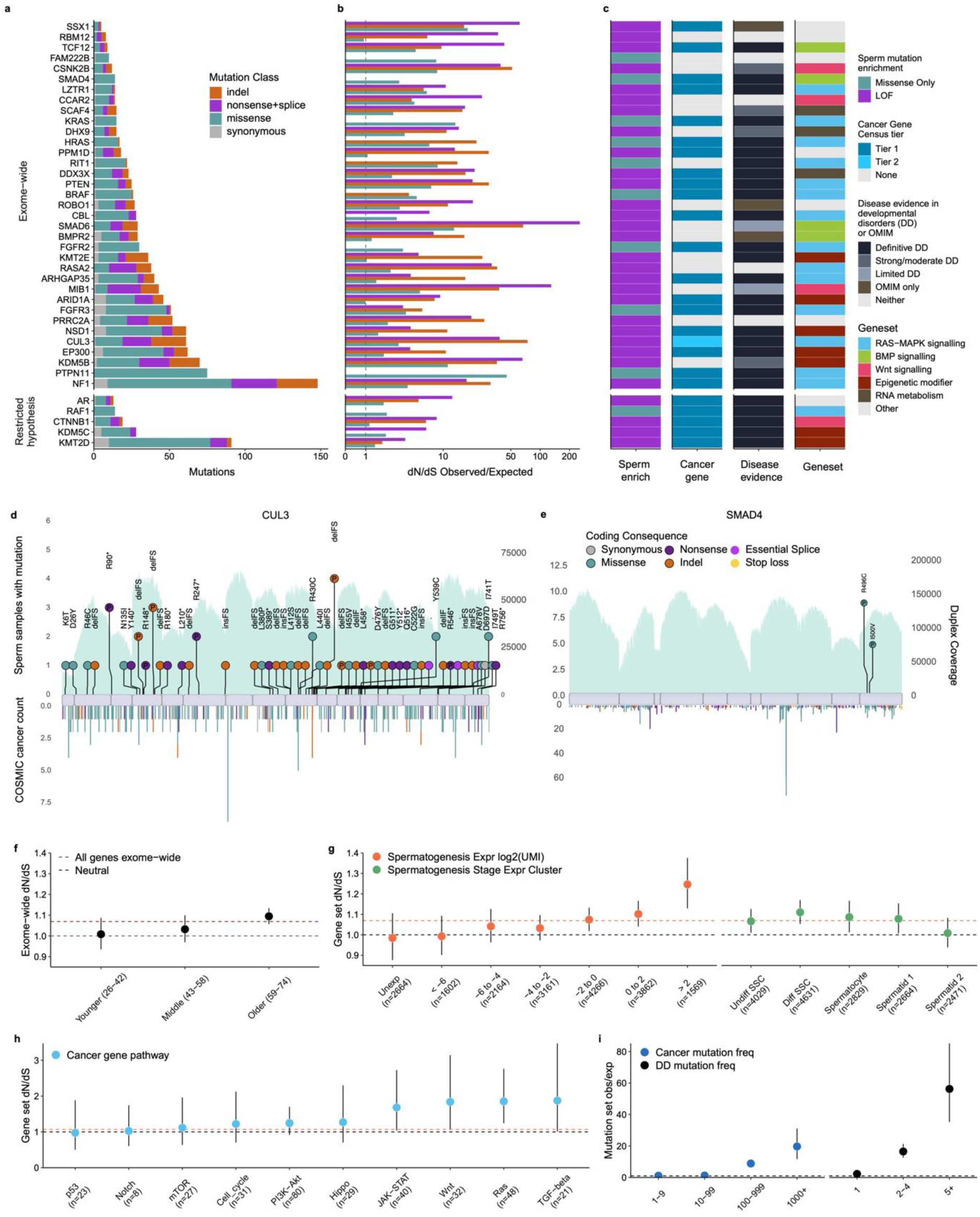
Germline positive selection. **a,b,c** Genes with significant dN/dS ratios from exome-wide and restricted hypothesis tests. **a,** Mutation count split by mutation class. **b,** Enrichment over expectation of mutation classes. **c,** Mutation type driving dN/dS enrichment, COSMIC^32^ cancer gene tier, developmental disorder gene link in DDG2P^33^, and potential germline selection geneset. **d,e,** Observed sperm mutations across the cohort for *CUL3* and *SMAD4* where the height of the “lollipop” represents the number of unique samples with a mutation at that location and the colour represents its mutation type. Mutations are labelled with their amino acid consequence for point substitutions or their insertion (ins)/deletion (del) consequence of in frame (IF) or frameshift (FS). A “P” indicates that the variant is classified as pathogenic/likely pathogenic in ClinVar^34^. Exons are shown as purple rectangles and the blue background represents the total duplex coverage across the cohort. Lines below the gene indicate COSMIC somatic mutations in cancer within that gene^32^. **f-h,** dN/dS ratios for sperm SNVs across sets of individuals or genes, where the dotted black line indicates neutrality and the dotted orange line represents the cohort average across all genes. **f**, Exome-wide dN/dS ratios in sperm for the cohort split into thirds by age. **g,** Expression gene sets from single-cell sequencing of germ cells^31^. Expression levels represent 7 bins of mean expression levels across germ cell stages and expression clusters represent genes most characteristic to certain germ cell stages. Germ cell types include undifferentiated and differentiated spermatogonial stem cells (SSCs), spermatocytes, round spermatids (1) and elongating spermatids (2). **h,** Germline selection genes and cancer gene census genes split by ten canonical cancer pathways in KEGG^35^. **i,** The observed/expected mutation rate in sperm for bins of mutations. COSMIC and DDD are bins of variants that have been seen at different levels of recurrence. Error bars depict 95% CIs.

We next compared the dN/dS ratios across gene sets related to spermatogenesis expression^31^ (**Fig. 2g**). We find that the gene sets with the highest dN/dS ratios are those which are highly expressed during spermatogenesis (1.25, 95% CI 1.13-1.38) and most specific to differentiated spermatogonial stem cells (1.11, 95% CI 1.05-1.17). In contrast, the genes which are unexpressed in spermatogenesis (0.98, 95% CI 0.88-1.11) and the genes most specific to elongating spermatids (1.01, 95% CI 0.94-1.08) show dN/dS ratios close to neutrality. These results are consistent with the understanding that excess nonsynonymous mutations observed in sperm confer a competitive advantage earlier in their cell lineage, specifically in the spermatogonial stem cells of the testis^15^.

### Discovery of novel genes and pathways under positive selection in the germline

We then investigated which genes were driving the signal of positive selection using the combination of the exome and targeted panel datasets (**Methods**). We applied dN/dS tests for excess non- synonymous mutations at both the gene-wide and SNV hotspot levels, which together identified 40 genes under significant positive selection. Of these, 35 genes reached exome wide significance at the gene level (FDR q < 0.1; **Supplementary Table 3**) and/or contained one of 17 exome-wide significant hotspots (q < 0.1; **Extended Data Table 1**). The genes *PTPN11, MIB1, RIT1, FGFR3, EP300,* and *FGFR2,* were significant in both the gene and hotspot tests, *KDM5B, NF1, SMAD6, CUL3, RASA2, PRRC2A, PTEN, ROBO1, DDX3X, CSNK2B, KRAS, PPM1D, ARID1A, BRAF, HRAS, KMT2E, SCAF4, BMPR2, TCF12, CCAR2, DHX9, NSD1, LZTR1, ARHGAP35, CBL, SSX1,* and *RBM12* were significant in only the gene test, and *SMAD4* and *FAM222B* were significant in only the hotspot test (**Fig. 2a,b**). We excluded the major seminal fluid component gene *SEMG1* from this list, despite excess indels driving gene-level significance. This gene is expressed at extremely high levels in seminal vesicles and unexpressed during spermatogenesis^36^, suggesting that the enrichment may be the result of a known process of indel hypermutation in highly-expressed genes^37,38^ from a small contamination of seminal vesicle DNA, rather than selection in germ cells.

Subsequently, we carried out restricted hypothesis dN/dS tests at the per-gene and per-site level. The gene level test examined only the set of 263 canonical cancer driver genes on our target panel and the site level test used a set of 1,963 sites composed of known cancer hotspots and recurrent DNM sites from the DDD cohort^39^. This identified 5 additional genes: *KDM5C, KMT2D, AR, CTNNB1,* and *RAF1* and 7 hotspots not already significant at the exome wide level (q < 0.1; **Fig. 2a,b**; **Supplementary Table 4; Extended Data Table 1**).

Genes linked to germline positive selection to date all operate through activating missense mutations, with 12 linked to the RAS-MAPK signalling pathway^18^ and one (*SMAD4*) linked to TGF-β/BMP signalling^40^. Our findings replicate *SMAD4* and 8 of the 12 RAS-MAPK pathway genes as under significant positive selection in this dataset. The 4 genes which did not reach significance (*MAP2K1, MAP2K2, SOS1,* and *RET*) each had between 2- and 4-fold enrichment of missense mutations, which corresponded to nominally significant missense enrichment in all 4 genes (p < 0.1). Given the direct evidence for these genes driving clonal selection in testis and nominal enrichment from sperm sequencing, we expect that each will reach exome-wide significance with deeper sequencing.

We estimate that together, the 44 genes linked to germline selection here or in previous studies, contain an estimated 357 (95% CI: 319–387) excess non-synonymous variants in exome sequenced samples. This would account for 23% (95% CI: 14%–43%) of the total estimated driver variants across the exome. The wide confidence intervals and the sensitivity of this estimate to the mutation model used (**Supplementary Note 3**) suggest that small uncertainties in mutation rates, when propagated across the exome, make it difficult to precisely estimate the fraction of drivers explained. Nevertheless, the findings suggest that additional driver genes remain to be discovered.

The 31 newly identified genes demonstrate that germline positive selection is not restricted to activating mutations or to the RAS-MAPK pathway. For instance, 30/31 of the novel genes are enriched for loss-function mutations such as nonsense, splice, and indel variants, suggestive of protein-inactivating mechanisms of selection (**Fig. 2c,d**). Splitting the germline selection genes and known cancer genes by ten canonical cancer pathways within the Kyoto Encyclopedia of Genes and Genomes (KEGG)^35^, we find that the top gene groups enriched in dN/dS are RAS-MAPK, Wnt and TGF-β/BMP signalling (**Fig. 2h**). Indeed, many of the novel selection genes are linked to the RAS-MAPK pathway, such as *NF1, CUL3* and *LZTR1*, Wnt signalling (i.e. *CSNK2B, MIB1, CCAR2*), and TGF-β/BMP signalling (i.e. *SMAD6*, *TCF12, BMPR2*) (**Fig. 2c**). We also identified a number of genes that encode epigenetic modifiers (i.e. *KDM5B, KDM5C, ARID1A, KMT2D, KMT2E, EP300, NSD1*), and genes encoding RNA metabolism proteins (i.e. *DHX9, DDX3X, SCAF4*). These findings highlight a new diversity of genes, mutational mechanisms, and pathways driving germline selection, however future work will be needed to confirm the specific pathways and roles through which these genes drive clonal expansion during spermatogenesis.

It has been observed that cancers and germline developmental disorders share causal pathways and genes^41–44^. Notably, the 13 genes previously linked to germline positive selection are all known cancer and known developmental disorders genes^18,40^. This pattern holds, but to a lesser extent, in the new germline selection genes identified here: 16 of 31 genes are tier 1 or 2 cancer census genes^32^ and 24 of 31 are linked to monogenic developmental disorders in the DDG2P database^33^ (**Fig. 2c; Supplementary Table 5**). Among the positively selected genes which are not associated with monogenic developmental disorders, three are linked to other monogenic disorders^45^ (**Fig. 2c; Supplementary Table 5**). These include *BMPR2* associated with pulmonary arterial hypertension^46–48^*, ROBO1* associated with pituitary stalk interruption syndrome^49,50^ and *SSX1* associated with X-linked spermatogenic failure^51^.

The overlap between germline positive selection genes and cancer/developmental disorders is also apparent at the variant level. Somatic mutations that are most frequently observed (>50 times) in the COSMIC cancer database are enriched 11-fold (95% CI: 6-20) among our sperm mutation dataset after adjusting for expected mutation rate (**Methods**). Similarly, germline mutations that are most frequently observed (>5 times) in a large cohort of children with developmental disorders are enriched 66-fold (95% CI: 41-100) in our sperm mutation dataset (**Fig. 2i**). In addition, the mutation types (e.g. missense or protein truncating variants) enriched in sperm for a given gene are largely consistent with those enriched in cancer and those causal for developmental disorders (**Extended Data Fig. 5**a-c). These results show a clear overlap between genes, hotspots, and mutation mechanisms which drive germline positive selection, cancer, and developmental disorders. A notable exception to this pattern is *SMAD4*, which has two distinct missense hotspots in sperm that are developmental disorder hotspots causal for Myhre syndrome^52^ but that are not often seen in cancers, replicating recent findings^40^ (**Fig. 2e**).

### Positive selection drives enrichment of disease-causing mutations in sperm

Given the association of many positively selected genes to disease, it is of interest to assess to what degree germline positive selection may increase the fraction of sperm carrying potential disease-causing mutations, and thus the birth prevalence of the associated disease. To estimate the fraction of sperm carrying specific classes of variants, we aggregated the variant allele frequencies (VAFs) of different mutation types and compared this to expected values. Expected values were generated using the custom fit *dNdScv* mutation model (adjusted for base pair coverage, CpG methylation, and trinucleotide context) and normalised to account for the linear impact of age on mutation rate (**Methods**).

We find that the fraction of sperm carrying a non-coding or synonymous mutation increases linearly with age as predicted by the model (**Extended Data Fig. 6**). In contrast, the frequency of missense, truncating (nonsense and splice) variants, and coding indels deviate above expected values in older individuals, consistent with dN/dS results and indicative of positive selection acting over time (**Extended Data Fig. 6**).

We then generated a list of likely monoallelic disease-causing mutations, which includes ClinVar^34^ pathogenic/likely pathogenic variants and highly damaging variants in high confidence monoallelic developmental disorder genes from DDG2P^33^ (loss-of-function or missense CADD^53^ score >30). This list represents a conservative estimate of disease-causing mutations due to the incomplete discovery and annotation of disease-causing variants and genes. We found that the observed fraction of sperm containing disease mutations was markedly higher than that expected under a germline mutational model at all ages of our cohort. The expected fraction of sperm with a likely disease mutation ranged from 0.73% in 30-year-olds to 1.6% in 70 year olds, whereas, fitting a quasibinomial regression, the observed fraction of sperm with a likely disease mutation in each age bracket ranged from 2% (95% CI: 1.6%-2.5%) in 30-year-olds to 4.5% (95% CI: 3.9%-5.2%) in 70 year olds (**Fig. 3c**). These differences represent similar enrichments of 2.8-fold (95% CI: 2.2 to 3.5) and 2.9-fold (95% CI: 2.5 to 3.3) in 30- year-olds and 70-year-olds respectively.

**Figure 3.**
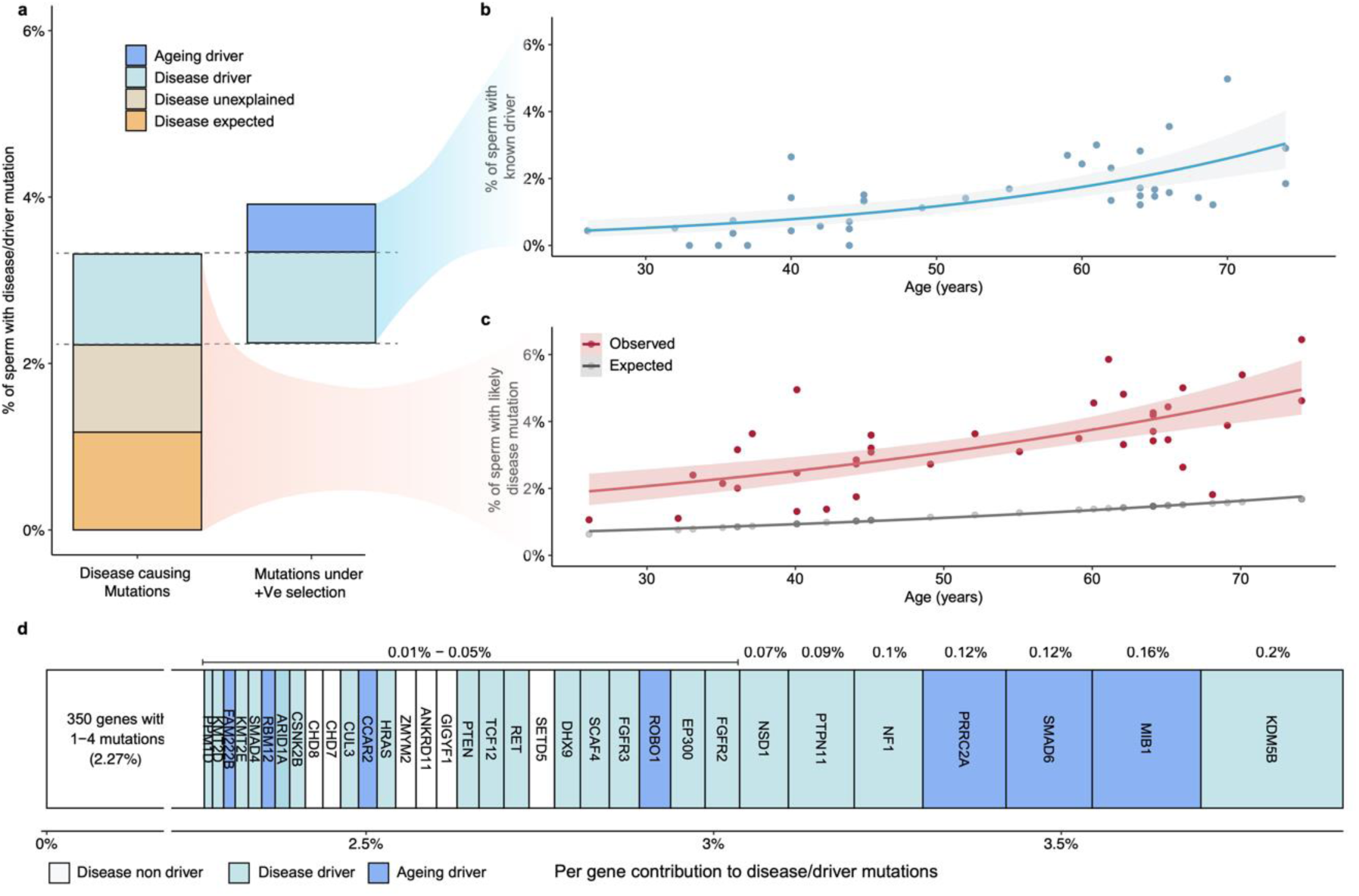
Pathogenic burden. **a,** Estimated mean percentage of sperm in the cohort carrying a likely monoallelic disease mutation (Left) or a driver mutation in a germline selection gene (Right). Disease mutations are divided into the fraction that was expected from the mutation model, the portion explained by driver variants and the portion unexplained. Driver mutations are split by those contributing to the disease mutations and the remainder, ‘Ageing drivers’. **b,** Estimated percentage of sperm per individual carrying a driver mutation by age. **c,** Observed and expected percentages of sperm with likely disease mutation by age. **b,c,** Model fits represent quasibinomial regressions with 95% confidence intervals. **d,** Cohort means from (**a**) split by gene and ordered by estimated mutation percentage. Per-gene contributions are shown above each gene; the summed contributions of all genes are shown below. Genes with 4 or fewer variants are grouped on the left with a condensed x-axis for clarity.

Interestingly, the disease cell fraction estimates are made up of many low frequency variants rather than being driven by individual high VAF mutations. The estimates in exome samples are made up of a mean of 18.3 unique variants (range 4-62) per individual. Furthermore, 692 of 696 (99.4%) of all those variants are only observed in a single sperm cell, similar to the average of all variants (99.5%).

We next investigated to what degree the observed enrichment of disease mutations can be attributed to driver mutations in positively selected genes. Fitting a quasibinomial regression, we observe a strong positive correlation between age and driver rate (*P* = 7.95e-06) with an estimated 0.5% (95% CI: 0.3%- 0.8%) of sperm from individuals at age 30 and 2.6% (95% CI: 2.0%-3.3%) of sperm from individuals aged 70 carrying a known driver mutation (**Fig. 3b**). However only about two thirds (65.6%) of those driver mutations meet our criteria of likely disease-causing.

Mutations in sperm that are likely disease-causing and those that are known driver mutations therefore represent overlapping but distinct annotations (**Fig. 3a**). Across the cohort, an estimated 3.3% of sperm carry a likely disease-causing mutation. Of this, approximately one-third (1.2%) is expected by the neutral mutation model, another third (1.1%) is explained by known driver mutations, and the remaining third (1.0%) is unexplained by either source. These findings suggest that the increase in likely disease-causing mutations is largely driven by germline positive selection, but also indicate that additional driver genes with disease associations remain to be identified.

Examining driver mutations which do not meet our criteria of a likely disease-causing mutation, we find that they impact an estimated 0.6% of sperm across the cohort. The consequences of these mutations are unclear. For instance, driver variants in *SMAD6*, which has a variably penetrant link to congenital phenotypes^54^, may be disease-causing in some cases but not others. Other potential consequences include mutations that are disease-causing but not yet annotated as such, less able to fertilise an egg, embryonic lethal, or biallelic disease-causing.

Much of the fraction of sperm with a disease and/or a driver mutation can be attributed to a small number of genes in the exome. From 374 genes with at least one such variant, the 33 genes with ≥5 independent mutations, most of which are under significant positive selection (26/33), represent 42.8% of the disease/selection fraction in sperm (**Fig. 3d**). Strikingly, 6 of those genes, all of which are under significant positive selection, (*KDM5B, MIB1, SMAD6, PRRC2A, NF1,* and *PTPN11*) together explain over 20% of the disease/selection fraction. This suggests that although individual mutations we observe are at low frequencies, positive selection systematically favours the likelihood of observing variants in driver genes.

We next sought to examine whether there are risk factors other than age which contribute to the accumulation disease or driver mutations in sperm. Currently known mutagenic effects in the male germline include chemotherapy and inherited DNA repair defects^39,55^ and small effect size influences of genetic ancestry and smoking^56^. While the cohort did not include any individuals with known chemotherapy treatments or DNA repair defects, phenotype data was available for BMI, smoking, and alcohol consumption, all of which have some evidence of driving mutation burden or driver mutation rate in some somatic tissues^57^. We used multivariate generalised linear models to test the association between these factors and measures of mutation burden, signatures, and driver cell fractions, correcting for multiple testing (**Methods; Extended Data** Fig. 6). Regardless of the sperm sequencing set examined (targeted, exome, or whole genome) only age was significantly correlated with measures of mutational burden, signatures, and driver cell fractions. No significant effects were found for BMI, smoking pack-years, or alcohol drink-years. However, in blood samples, age, smoking, and alcohol consumption showed significant effects on SNV and SBS5 burdens. These results suggest that, unlike many somatic tissues, the male germline mutation landscape may be largely protected from these risk factors, although larger cohorts will be needed to interrogate possible small effect sizes.

### Selection in germ cells relative to single generation and population-level variants

Mutations in sperm account for ∼80% of DNMs and are therefore also the origin of most population level variants. Comparisons between these different sources of germline variants provide an opportunity to explore how positive selection in the male germline may manifest over time.

Examining control DNMs from offspring without clinical phenotypes^58^ we found a dN/dS ratio consistent with neutrality of 0.98 (95% CI 0.90-1.08; **Fig. 4a**). In contrast, the dN/dS ratio in DNMs from offspring with developmental disorders showed a large enrichment of nonsynonymous variants, as previously reported^39^: 1.36 (95% CI 1.33-1.39; **Fig. 4a**). However, ascertainment biases in these cohorts suggest that these dN/dS ratios may not accurately reflect the dN/dS ratio of DNMs entering the population. Future large DNM studies of birth cohorts will likely be required to give less biased estimates.

**Figure 4.**
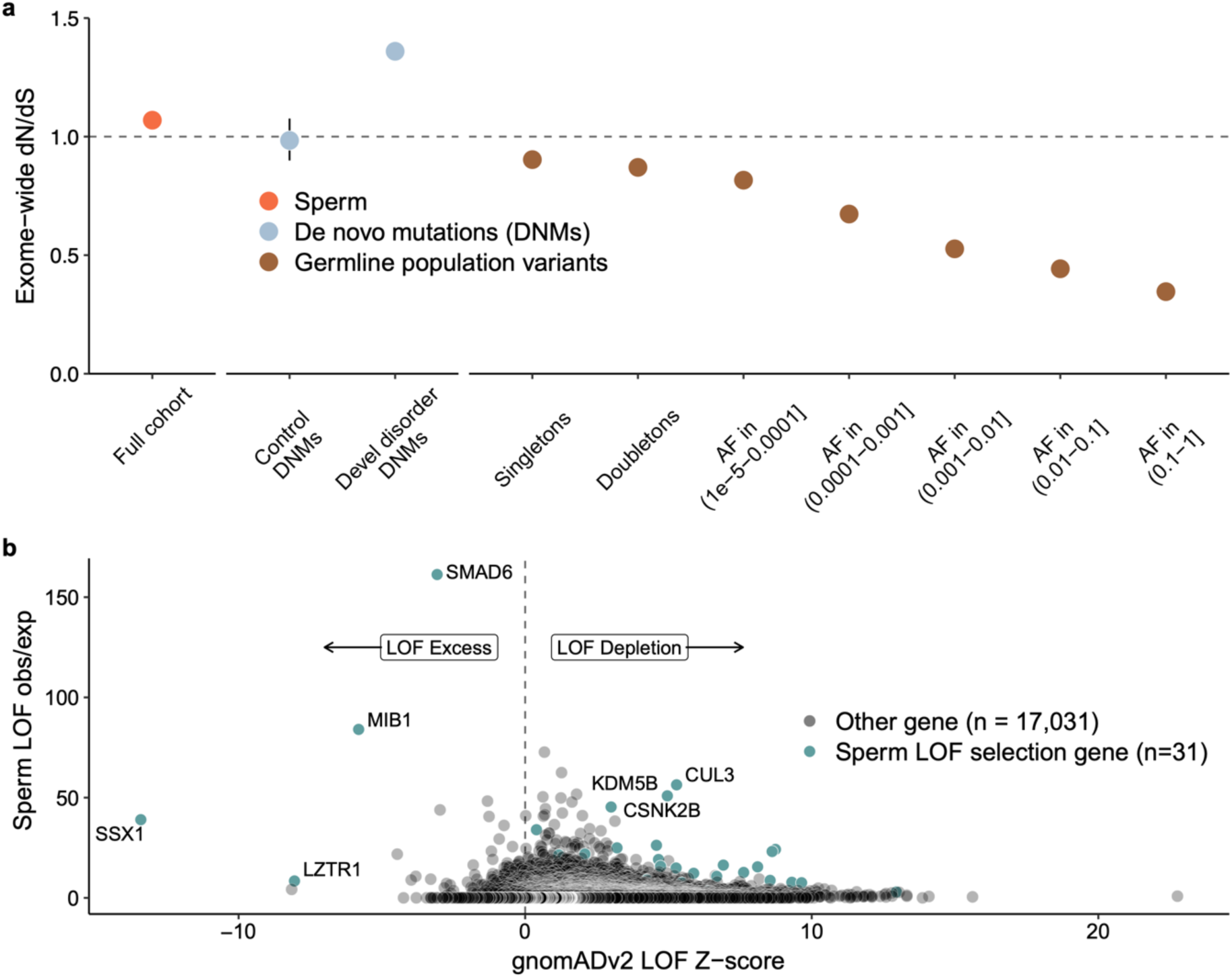
Comparison to population variation. **a,** Exome-wide dN/dS ratios across different variant sets, including sperm variants from all exome sequenced samples, de novo mutations (DNMs) from a collection of healthy trios^58^ and the Deciphering Developmental Disorders (DDD) cohort^39^; and population variants from gnomAD^59^ split by allele frequency (AF). **b**, Observed/expected enrichment of missense and loss-of-function (LOF; essential splice, nonsense, indel) variants in positively selected genes within sperm (x-axis) from dN/dS models vs gnomAD LOF z-score. Positive z-scores indicate LOF depletion, while negative ones indicate excess over expected. Error bars indicate 95% CIs.

The pattern of largely neutral dN/dS ratios in control DNMs contrasts with the strong evidence of negative selection in human single nucleotide polymorphisms (SNPs), particularly in common SNPs. To shed light on this, we calculated dN/dS ratios for population variants at different allele frequencies (AF) in the population using data from gnomAD^59^ (**Fig. 4a**). This revealed a decay in dN/dS ratios for population variants with higher AFs, a pattern consistent with purifying selection operating over many generations on germline SNPs. Altogether, these analyses suggest that the dominant selection force on germline mutations is positive selection during spermatogenesis but negative selection between generations. This dynamic mirrors the well-established contrast between selective forces on cancer mutations and those on germline mutations in populations^29^.

We then compared the per-gene enrichment of loss-of-function mutations in sperm to those of population germline variants using the gnomAD loss-of-function z-score. This z-score is a measure of how significantly the observed counts differ from expectations of a germline mutation model. The vast majority of genes in gnomAD have a positive z-score, indicative of depletion of LOFs and the negative selection expected in populations. Of the 31 significant germline selection genes with LOF enrichment in sperm (range 3-fold to 50-fold), 27 were depleted for LOFs in gnomAD. Each of these genes has a disease phenotype, consistent with a model by which these genes are selected for in spermatogenesis but purged from the population by strong negative selection. Interestingly, 4 significant germline selection genes had more loss-of-function mutations than expected in gnomAD: *SMAD6, MIB1, LZTR1,* and *SSX1* (**Fig. 4b**). The latter 3 of these genes are 3 of the 4 strongest outliers of LOF enrichment of all genes in gnomAD v2 and were given a cautionary outlier label for unexplained LOF enrichment. Our results suggest that the explanation behind their apparent enrichment in gnomAD is that germline positive selection introduces them at a higher rate than for other genes and negative selection against these variants is not strong enough to mask it.

## Discussion

We sequenced sperm and blood from healthy men spanning a wide age range to quantify mutation rate and positive selection in the male germline. The observed mutation rates and mutational signatures in sperm were consistent with those from family trio and testis sequencing studies^7,27,60–63^. We find that, despite sharing the same mutational signatures, SBS1 and SBS5, mutations accumulate at approximately an 8-fold lower rate in sperm compared to blood. This supports our previous observations comparing germline mutations in testes to a wide range of somatic tissues^7^ and emphasises the protected nature of the germline relative to the soma.

Analysing over 35,000 coding variants from sperm exome-wide, we build on the 13 previously identified germline selection genes, identifying an additional 31 genes under positive selection. This provides a foundational catalogue of genes under selection in the male germline and expands the diversity of pathways and mutational mechanisms linked to selection in this tissue.

Our findings have significant implications for studies relying on germline mutation models, as they do not currently account for germline positive selection. As shown here, this can lead to inaccurate constraint metrics in population cohorts. Importantly, this bias can also affect the identification of novel disease-causing genes from DNM enrichment tests. For instance, the recent identification of an excess of *de novo* loss-of-function mutations in *MIB1*^39^ likely reflects germline selection rather than disease association. Loss-of-function variants in *MIB1* are more common in population cohorts than expected for a gene associated with developmental disorders, and carrying one of these variants does not correlate with developmental disorder phenotypes^64^. In principle, adopting case-control tests for DNM enrichment should help to exclude genes under germline selection that are not linked to the disease. However, sufficiently large control trio datasets, as well as close age matching of controls to account for the age dependency of germline selection, will be needed to ensure sufficient statistical power. Until such resources become available, analyses of DNM enrichment in disease should take into account the evidence for germline selection influencing individual genes presented here (**Supplementary Table 3**).

Unlike the example of *MIB1*, most genes under positive selection during spermatogenesis are known to be associated with severe monogenic disorders with mutation mechanisms under positive selection matching those associated with disease. We demonstrate that this positive selection leads to a striking 2-3 fold enrichment in the fraction of sperm carrying a likely disease mutation across the age range studied. As a result, we estimate that 3-4% of sperm from men over 50 carry a likely disease-causing mutation. Somewhat reassuringly, we note that typical paternal ages at conception are younger than the average age of sperm donors studied here, and that the impact of germline positive selection will be correspondingly weaker. Future investigations, including sequencing of sperm from cohorts focused on men under the age of 30 will aid in developing estimates of germline selection strength in this age range.

A key consideration when interpreting the fraction of disease mutations in sperm is that this fraction may not directly correspond to the rate at which these variants are observed in live births. There are a number of reasons why, for some genes, the transmission rate to live births could be lower than those observed in sperm, including impaired ability of sperm to fertilise an egg, embryonic lethality, or increased pregnancy loss (**Supplementary Note 4**). Future studies, such as sequencing of trio DNMs from large birth cohorts, will be needed to quantify the relationship between the rate of positively selected disease mutations in sperm and disease incidence in populations.

While up to 3-5% of sperm in middle-aged to elderly individuals harbouring a known driver mutation has a large impact on offspring disease risk, it is on the low end of the spectrum of estimates in proliferative somatic tissues. For instance, in comparable age groups, more than 40% of cells in endometrial and esophageal epithelium carry a driver mutation^2,5^, whereas only a few percent of cells are estimated to carry a driver mutation in healthy colon or liver^3,4^. For blood, on average only a few percent of cells carry a driver mutation in middle aged individuals, but some individuals can present large clonal expansions due lack of severe spatial constraints for clonal expansion^9^. While the germline mutation rate is under evolutionary pressure to remain low to prevent detrimental mutations across the genome, it is perhaps under particularly strong pressure to keep driver mutation rate low, as a single germline driver mutation can cause disease in offspring. It is likely that the tubular organisation of the testis provides strong spatial constraints to prevent large clonal expansions, limiting the accumulation of driver mutations in sperm despite the large number of cell divisions required to sustain sperm production. The low relative rate of driver mutations in the male germline is also consistent with the unique aspects of spermatocytic tumours, the tumour type thought to derive from spermatogonial stem cells in the testis. Spermatocytic tumours are rare relative to most somatic tumours and are primarily driven by chromosome aneuploidies^65^ rather than the classical sequential acquisition of driver mutations leading to cancer transformation^66,67^ observed in driver rich somatic tissues.

The findings of this study provide important insights into the historically underexplored reproductive ageing risks associated with the male germline. This contrasts with the well-established relationship between maternal ageing and reproductive risks, where decreasing oocyte quality with age leads to increased rates of pregnancy loss and aneuploidy^68^. Our results demonstrate that driver mutation accumulation from the male germline’s continuous cell proliferation is a substantial risk, though one spread across many genes. As trends toward delayed reproduction continue^69^, it is essential to recognise that both paternal and maternal ageing contribute to elevated risks for offspring, albeit primarily through different biological mechanisms. Future research will refine our understanding of selective pressures and disease risk associated with germline mutation, enhancing our understanding of their implications for human reproduction and health.

## Methods

### Ethics

This study was carried out under TwinsUK BioBank ethics, approved by North West – Liverpool Central Research Ethics Committee (REC reference 19/NW/0187), IRAS ID 258513 and earlier approvals granted to TwinsUK by the St Thomas’ Hospital Research Ethics Committee, later London – Westminster Research Ethics Committee (REC reference EC04/015).

### Sample collection

Bulk semen samples were collected or obtained from archival samples with informed consent from 75 research participants within the TwinsUK cohort^26^. Archival whole-blood samples were also obtained from 67 of those men from the TwinsUK BioBank. A total of 104 semen samples spanned an age range of 24-75 years and included 29 men with 2 timepoints separated by a mean of 12.1 years (range 12-13 years) and the remaining 46 men with a single timepoint. A total of 133 blood samples were collected at an age range of 22-83 years. There were 11 men with a single blood timepoint, 47 with two timepoints, 8 with three timepoints and 1 with four timepoints. The mean interval between blood timepoints was 8.1 years (range 1-13 years). Within the cohort there were a total of 9 monozygotic (MZ) twins and 3 dizygotic (DZ) twin pairs. Counts of samples, timepoints, and twin pairs which were successfully sequenced and passed analysis quality control thresholds are summarised in **Supplementary Table 1**.

### Metadata

Metadata for self-reported age, height, weight, ethnicity, twin zygosity, smoking and alcohol consumption were obtained from questionnaires provided by TwinsUK taken periodically. All individuals that provided ethnicity information indicated “white”. BMI was calculated as weight/height^2^. A smoking pack year was defined as 365 packs of cigarettes and total pack years was calculated using the highest estimate across all questionnaires from cigarettes per day or week and total years smoked. Alcohol drink years was calculated from using average weekly alcohol consumption extrapolated to the duration of adult life before sampling (age - 18).

### DNA extraction

DNA was extracted from sperm samples using the Qiagen QIAamp DNA Blood Mini Kit. Isolation of genomic DNA from sperm; protocol 1 (QA03 Jul-10) was followed with the exceptions of substituting DTT in place of β-mercaptoethanol for Buffer 2 and substituting Buffer EB in place of Buffer AE for elution of DNA.

DNA was extracted from whole blood using the Gentra Puregene Blood Kit, following the protocol for 10ml of compromised whole blood from the Gentra Puregene Handbook version 06/2011.

### Targeted gene panels

Three separate Twist Bioscience gene panels were used for targeted NanoSeq sequencing in this study: 1) a custom pilot panel of 210 genes; 2) a similar but extended custom panel of 263 genes (**Supplementary Table 2**); 3) a default exome-wide panel of 18,800 genes. The two custom gene panels are highly similar with the extended panel being almost exclusively regions added to the pilot panel. From the 84 samples that underwent targeted sequencing, 13 were sequenced using a pilot panel of 210 canonical cancer/somatic driver genes, and all 84 were sequenced using the extended panel of 263 genes. Sequencing coverage and mutations were merged from samples sequenced on both targeted panels. The custom panels were designed by gathering sets of published lists of genes implicated as drivers in cancers^71–74^ and somatic tissues^1,75^ as described in (Lawson A.R., Abascal F., P.A. Nicola et al., manuscript submitted for publication).

### Sequencing and preprocessing of NanoSeq libraries

Restriction-enzyme whole genome NanoSeq libraries were prepared as described in Abascal et al.^23^ and subjected to whole genome sequencing at target 20-30x coverage on NovaSeq (Illumina) platforms to generate 150-bp paired-end reads with 9-10 samples per lane. Standard whole genome sequencing of blood (31.7x median coverage) was used to generate matched-normal libraries for both restriction-enzyme NanoSeq blood and sperm.

Targeted and exome NanoSeq libraries were prepared via sonication and 1-2 rounds of pull down of target sequences as described in (Lawson A.R., Abascal F., P.A. Nicola et al., manuscript submitted for publication). They were then sequenced with NovaSeq (Illumina) platforms to generate 150-bp paired-end reads with 7-8 samples per lane for the targeted panel and 2 lanes per sample for the exome panel.

### Base calling and filtering

All samples were processed using a Nextflow implementation of the NanoSeq calling pipeline (https://github.com/cancerit/NanoSeq). BWA-MEM^76^ was used to align all sequences to the human reference genome (NCBI build37). Restriction-enzyme NanoSeq samples were called with their matched WGS normal and default parameters except for var_b (minimum matched normal reads per strand) of 5 as needed for WGS normals, cov_Q (minimum mapQ to include a duplex read) of 15 and var_n (maximum number of mismatches) of 2.

For targeted and exome samples we leveraged the high sequencing depth and high polyclonality to exclude variants with VAF > 10% instead of using a matched normal. Default parameters of the calling pipeline except for cov_Q of 30, var_n of 2, var_z (minimum normal coverage) of 25, var_a (minimum AS-XS) of 10, var_v (maximum normal variant allele frequency (VAF)) of 0.1, and indel_v (maximum normal VAF) of 0.1. Post variant calling, we further filtered variants to those below 1% VAF and those below 10% duplex VAF as variants above these cutoffs were highly enriched for mapping artefacts, particularly for indels. No excluded variants from these additional VAF thresholds were found to be likely driver or ClinVar pathogenic variants; all exclusions were inspected to confirm this.

A set of common germline variants from dbsnp^77^ and a custom set of known artifactual call sites in NanoSeq datasets were masked for coverage and variant calls as previously described^23^.

### Assessing DNA contamination

The single-molecule accuracy of the duplex sequencing method NanoSeq allows sequencing of polyclonal cell types such as sperm, but also renders mutation calls sensitive to a) non-target cell-type contamination and b) contamination of foreign DNA. Non-target cell-type contamination was evaluated using manual cell counting of semen samples, resulting in the exclusion of 6 samples with sperm count < 1 million sperm/mL. Sperm counting methods and analysis are detailed in **Supplementary Note 1**.

Foreign DNA contamination in whole genome NanoSeq samples was assessed using verifyBamID^78^, which checks whether reads in a BAM file match previous genotypes for a specific sample, with higher values indicating more contamination. Three blood whole genome NanoSeq samples were excluded based on a verifyBamID alpha value above the suggested cutoff of 0.005^23^. In sperm, we found that several samples had outlier mutation burdens with verifyBamID values just below the 0.005 cutoff. This is logical, as sperm has a much lower mutation rate compared to somatic tissues, for which the recommended cutoff was designed. Consequently, sperm samples are more sensitive to low levels of contamination. To account for this, we adjusted the verifyBamID alpha threshold for sperm to a more stringent level of 0.002, resulting in the exclusion of 3 samples on this criterion.

When assessing foreign DNA contamination in targeted and exome samples we found that 9 targeted and 3 exome samples had verifyBamID values above > 0.002, slight outlier mutation burdens, and high ratio of SNP masked variants to passed variants (4-fold to 16-fold more masked variants). Upon further investigation we found that all samples exceeding verifyBamID thresholds were processed in the same sequencing batch and that this contamination could be explained by inherited germline variants of other samples within that same batch. This suggests that a small amount of cross-contamination may have occurred during sample preparation or sequencing steps. In order to remove contaminant germline mutations we performed an *in silico* decontamination as previously described^23^. This involved calling germline variants from all targeted and exome samples using bcftools mpileup^79^ at sites where there were >10 reads and a mutation call with VAF > 0.3. All such sites were subsequently masked across all samples for both mutation calls and coverage, essentially extending the default common SNP mask to also include rare inherited variants across the cohort. This resulted in all samples previously identified as contaminated having mutation burdens consistent with their age and all having a ratio of masked to passed variants <0.1, and were thus retained for analysis.

### Corrected mutation burdens

Given that mutation rates are strongly influenced by trinucleotide composition, it is important to consider differences in sequence composition when comparing mutation rates in datasets that target different regions of the genome. For instance, it is known that coding regions such as those in NanoSeq target panels, are biased towards a higher mutation rate partially due to a higher GC density than non- coding regions^80^ which make up the majority of sequenced regions in whole genome NanoSeq datasets. To correct for this effect, in each sperm NanoSeq dataset we generated a corrected mutation burden relative to the full genome trinucleotide frequencies as described previously^23^.

### Comparison of NanoSeq and WGS burden estimates

In order to compare whole genome NanoSeq mutation burdens to mutation burden from standard whole genome sequencing (WGS) we multiplied the corrected mutation burden estimates described in the previous section by the genome size per cell type. We assumed 2,861,326,455 mappable base pairs in a haploid cell for germline datasets and the diploid equivalent of 5,722,652,910 base pairs for blood.

External datasets for comparison to NanoSeq results were processed in order to achieve comparable burden estimates. For testis WGS samples^70^ we implemented the method described in Abascal et al.^23^ that restricts analysis to regions with high coverage (20+ reads) that overlap with NanoSeq covered regions. Additionally, we corrected for differences in trinucleotide background frequencies relative to the full genome as described in the previous section. For trio paternally phased DNMs from standard sequencing, as a callable genome size per sample following thorough filtering was available, we generated the mutation per cell estimate by multiplying the paternally phased DNM count by the ratio of the sample’s callable genome to total genome size. For comparison to cell types in blood we compared directly to the published mutation burden regressions^8^.

### Mutation burden regressions

To investigate the relationship between age and mutation burdens, we performed linear mixed-effects regression analyses. For each tissue and mutation type where a regression was performed, the model was constructed using the ‘lmer’ function from the ‘lme4’ package^81^ in R. Each model included age at sampling as a fixed effect and a random slope for each individual to account for multiple timepoint samples, specified as:

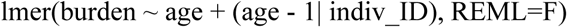

The 95% confidence intervals (CIs) for regression lines were calculated through bootstrapping by simulating prediction intervals. For each model, we generated 1000 bootstrap samples. Predictions and their associated standard errors were calculated for a sequence of ages from 14 to 84 years. The 95% CIs were then derived by determining the range within which 95% of the bootstrap sample predictions fell.

### Mutational signature analysis

We extracted *de novo* mutational signatures using Hierarchical Dirichlet Process (HDP; https://github.com/nicolaroberts/hdp), which is based on the Bayesian Hierarchical Dirichlet process. HDP was run with double hierarchy: (1) individual ID and (2) tissue types (either blood or sperm), and without the Catalogue Of Somatic Mutations In Cancer (COSMIC) reference signatures^82^ (v3.3) as priors, on the mutation matrices. The number of mutations were normalised for the tri-nucleotide context abundance specific for each sample relative to the full genome. Both clustering hyper- parameters, beta and alpha, were set to one. The Gibbs samples ran for 30,000 burn-in iterations (parameter “burnin”), with a spacing of 200 iterations (parameter “space”), from which 100 iterations were collected (parameter “n”). After each Gibbs iteration, three iterations of concentration parameters were conducted (parameter “cpiter”). Two components were extracted as *de novo* mutational signatures which were further reconstructed and decomposed into known COSMICv3.3 SBS signatures using SigProfilerAssignment (https://github.com/AlexandrovLab/SigProfilerAssignment). As a result, three COSMIC signatures: SBS1, SBS5, and SBS19, were reported.

### Quantifying selection with dN/dS

To examine genes under positive selection and quantify global selection we used the *dNdScv* algorithm^29^. This algorithm was extended using base pair level duplex coverage, methylation level and pentanucleotide context to capture more complex context dependent mutational biases, and to achieve more accuracy for our selection analysis. Detailed methods for input mutations, model selection and evaluation, site dN/dS tests, driver mutation estimation, and gene set enrichment are described in **Supplementary Note 3**.

### Gene disease and mechanism annotation

Positively selected genes were annotated with monoallelic disease consequences using the 2024-02-29 release of the Development Disorder Genotype - Phenotype Database (DDG2P)^33^ and the 2024-06-21 release of the Online Mendelian Inheritance in Man (OMIM) database^45^. OMIM annotations related to somatic disease, complex disease, or tentative disease associations were excluded.

Genes were also annotated for their potential mutation mechanism observed in sperm and cancer/developmental disorders when available. In sperm, genes were labelled as loss-of-function if they had nominal enrichment of nonsense+splice variants and/or indel variants (ptrunc_cv < 0.1 | pind_cv < 0.1) and 2+ loss-of-function mutations. There were two exceptions to this where genes met these thresholds but were labelled as activating due to having a restricted repertoire of loss of function mutations that are known to be oncogenic in cancers: *CBL* (LOFs in and downstream of the RING zinc finger domain)^83^ and *PPM1D* (LOFs in final two exons)^84^. All other genes had missense enrichment only and were labelled as activating. The mechanism in cancer was defined by using the ’Role in Cancer’ field of the COSMIC cancer gene census v99^32^ where ‘oncogene’ was labelled as activating and ‘tumour suppressor gene’ as loss of function. Annotations of a fusion mechanism were not displayed except for genes which had neither an oncogene, nor a tumour suppressor annotation which were labelled as ‘fusion only’. The developmental disorder mechanism was defined by using the variation consequence field of DDG2P where ‘restricted repertoire of mutations;activating’ was labelled as activating and ‘loss_of_function_variant’ was labelled as loss of function.

### Gene mutation plots

The “lollipop” gene mutation plots were created with a coordinate system where the 1 was the first coding base of the GRCh37-GencodeV18+Appris^85^ transcript of the gene. The data sources included protein domains from UniProt^86^, somatic mutations from the exome and genome wide screens of the COSMIC^32^ (v99), ClinVar release 2024.07.01^34^ pathogenic annotation, per base pair cohort wide (targeted + exome) NanoSeq coverage, sperm mutation count (number of independent individuals with a mutation) and mutation consequence and amino acid change annotated by the dNdScv algorithm^29^. These data were plotted with code modified from the lolliplot function in the trackViewer R package^87^.

### Variant annotation

Variants were annotated using Ensembl’s Variant Effect Predictor (VEP)^88^ with added custom annotations of mutation context, ClinVar release 2024.07.01^34^, Combined Annotation Dependent Depletion (CADD) version GRCh37-v1.6^53^ and average methylation level in testis. Methylation data was obtained from whole genome shotgun bisulfite sequencing methylation data from a 37 year old (ENCFF638QVP) and 54 year old (ENCFF715DMX) male testis from the ENCODE project^70^. The average methylation level was calculated by selecting CpG sites with coverage of 3 or more and averaging the percent of sites methylated between the two samples.

Variants were annotated as likely monoallelic disease-causing mutations if they met at least one of two criteria: 1) Reported in ClinVar as pathogenic, likely pathogenic, or if they were reported as ‘Conflicting_classifications_of_pathogenicity’ where the conflict was between reports of pathogenic/likely pathogenic and ‘Uncertain_significance’ with no reports of benign or likely benign and not specified as a recessive condition or 2) Were a highly damaging variant in a high confidence monoallelic developmental disorder genes from DDG2P^33^. Genes met criteria of a) allelic requirement being monoallelic_autosomal, monoallelic_X_hem, monoallelic_X_het, or mitochondrial, b) confidence in strong, definitive, or moderate and c) a mutation consequence of ‘absent gene product’. Highly damaging was defined as being a ‘HIGH’ impact variant in VEP annotation (frameshift splice_acceptor, splice_donor, start_lost, stop_gained, or stop_lost) or a missense variant with CADD^53^ score >30 (top 0.1% damaging).

Variants were defined as a likely driver if they met the ‘highly damaging’ criteria defined above in a significant germline selection gene with loss-of-function mutation enrichment or if they were in one of the 24 significant mutation hotspots. This resulted in 320 variants being labelled as likely drivers in exome samples.

### Cell fraction mutation estimates

To calculate the mean count of synonymous, missense or loss-of-function, or pathogenic mutations per sperm cell we summed the duplex VAFs of all variants in that class. For example, if an individual had three synonymous mutations each observed once with a duplex coverage of 100 at each of those sites, each of those variants would have a duplex VAF of 1/100 = 0.01. The sum of VAFs in this example would then be 0.03 and this would then be reported as the estimate for the mean count of synonymous variants per sperm cell for that individual. At low fractions such as 0.03, the mean count per cell is approximately equivalent to the percentage of sperm with this mutation class (3%) and thus the driver and disease mutations are reported as percentage estimates. At higher fractions (e.g. mean count > 1) the fractions are not equivalent to percentage as many cells will have multiple variants of that class and thus the estimates are reported as mean count.

Expected mean counts for SNVs were generated by annotating each possible substitution at each covered site with an expected number of mutations per sample as given by expCountSNV = context_mut_rate*duplex_coverage*age_correction. The context_mut_rate was given by the 208 basePairCov + cpgMeth trinucleotide mutation model estimates for that trinucleotide+methylation mutation context (**Supplementary Note 3; Supplementary Table 6**). Duplex coverage is the exact duplex coverage at that site for that sample. The age_correction parameter was given to normalise the mutation model estimates (derived from all exome samples) to the mutation rate of that sample based on age. Specifically, we fit a linear model to the mutation burden vs age of the exome samples and used this to generate a predicted mutation rate for each sample based on age. The per sample corrected parameter was calculated as the age predicted mutation burden divided by the mean mutation rate of all exome samples (3.89e-08). The resulting corrections spanned from 0.50 (youngest sample) to 1.42 (oldest samples). The expected indel mutation rate was calculated in the same way, except with a single mutation rate parameter (indels/bp) expCountIndel = indel_mut_rate*duplex_coverage*age_correction. The expected mean count was then calculated for each category (e.g. synonymous, likely disease) by summing the expected values for each SNV and/or indel base pair matching the relevant annotation. As background for possible ClinVar pathogenic variants we only considered indels of size 21 bp or less, the largest detected indel in the dataset. Regressions were fit with either linear models or generalised linear models (*glm* in R) with family = quasibinomial.

### Regression analysis

We tested for associations between mutation outcome variables from sperm genome, sperm exome, sperm targeted and blood genome NanoSeq data and the phenotype predictor variables of BMI, smoking pack years, and alcohol drink years. These tests were performed using a gaussian family generalised linear regression in R. For each mutation outcome variable the test took the form of: glm(mutationOutcome ∼ age_at_sampling + BMI + pack_years + drinkYears, family = “gaussian”).

The mutation outcome variables examined were SNV and indel burden from all 4 sequencing datasets, SBS1 and SBS5 count from sperm genomes, SBS1, SBS5, and SBS19 from blood genomes, and likely disease cell fraction and likely driver cell fraction from sperm targeted and sperm exomes. The significance of each predictor was assessed from the model’s summary output coefficients, and p-values were adjusted for 68 total tests using the false discovery rate method.

## Data availability

Raw sequencing data are available on the European Genome–Phenome Archive under accession number X. Additional individual-level data are not permitted to be publicly shared or deposited due to the original consent given at the time of data collection, where access to these data is subject to governance oversight. All data access requests are overseen by the TwinsUK Resource Executive Committee (TREC). For information on access to these genotype and phenotype data and how to apply, see https://twinsuk.ac.uk/resources-for-researchers/access-our-data/.

## Code availability

All scripts are available on github at https://github.com/mattnev17/spermPositiveSelectionManuscript.

## Data Availability

Individual-level data are not permitted to be publicly shared or deposited due to the original consent given at the time of data collection, where access to these data is subject to governance oversight. All data access requests are overseen by the TwinsUK Resource Executive Committee (TREC). For information on access to these genotype and phenotype data and how to apply, see https://twinsuk.ac.uk/resources-for-researchers/access-our-data/.

## Acknowledgements

We thank the TwinsUK research volunteers for participating in the study. We are grateful to Laura O’Neill, Calli Latimer and all of the CASM Support team at the Wellcome Sanger Institute for their assistance. We thank Charlotte Seymour, Elisa Ferraro, and Chris White from Cambridge IVF for training and guidance on sperm counting.

## Author Contributions

M.D.C.N., M.E.H. and R.R. wrote the manuscript; all authors reviewed and edited the manuscript. M.E.H., and R.R. supervised the project. M.P.G., S.W., K.S., and R.R. led sample procurement. M.D.C.N., T.B., and P.A.N. conducted sperm counting. A.R.J.L., P.J.C., K.R., S.V.L., S.W., and I.M. contributed to sample sequencing implementation. M.D.C.N., led the analysis of the data with help from A.R.J.L., R.S., F.A., M.H.P., A.C., P.A.N., A.B.O., R.E.A., M.R.S., P.J.C., I.M., M.E.H., and R.R.

---

**Extended Data Fig. 1.**
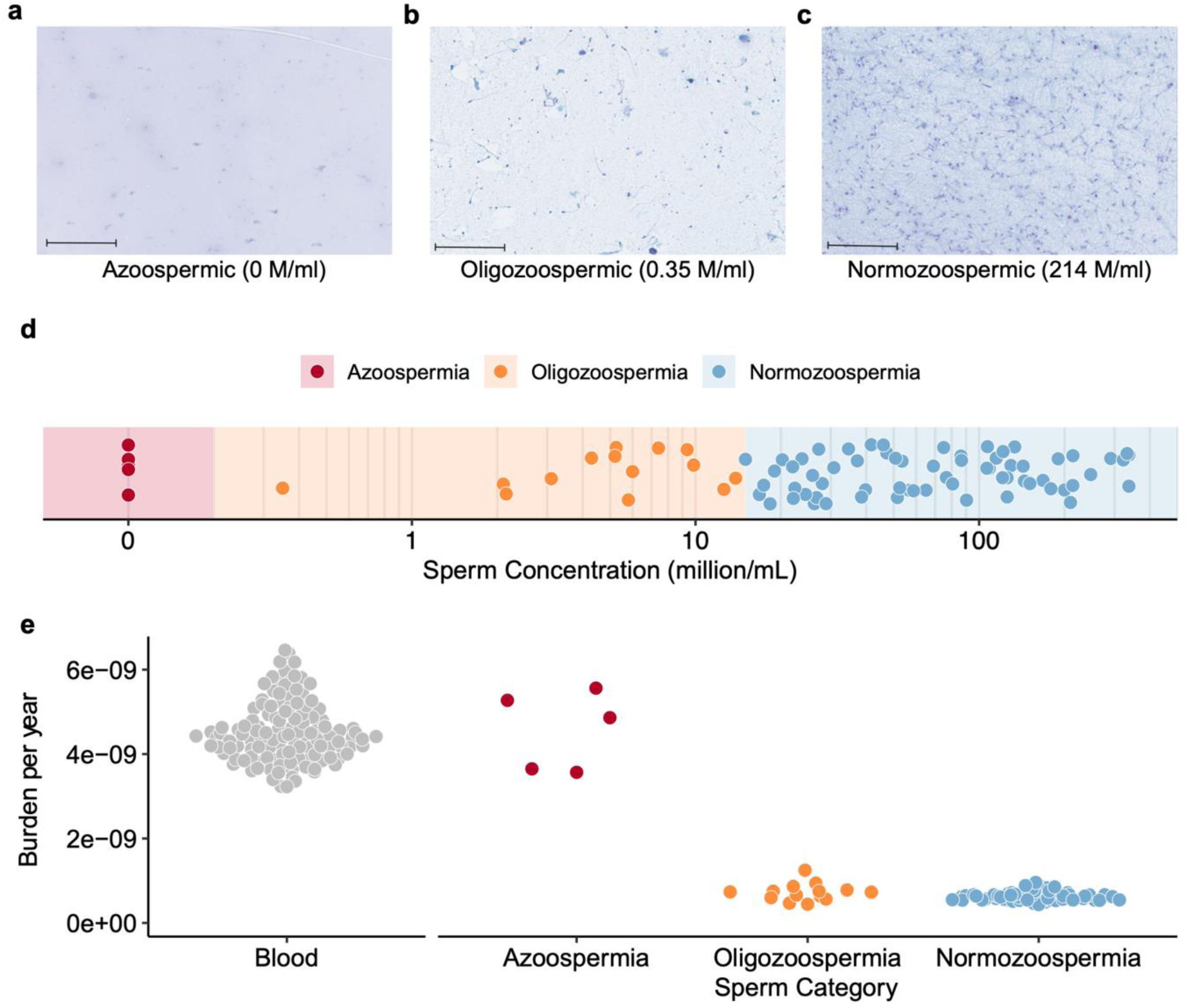
Sperm counting. **a,b,c,** Slides of Papanicolaou stained semen samples for (**a**) an azoospermic sample where no sperm cells are visible, (**b**) an oligozoospermic sample where a small number of sperm samples are visible and (**c**) a normozoospermic sample where many sperm cells are visible. Sperm concentrations are given for each sample in millions of sperm per ml (M/ml). The black band in the bottom left of each slide photo corresponds to 100µm. **d,** The distribution of sperm counts on a log scale among semen samples analysed with colour bands indicating the concentration bin of the sample. All samples below 1 million/mL were subsequently excluded. **e,** The distributions of mutation burden per year from blood samples and three categories of sperm samples broken down by sperm concentration.

**Extended Data Fig. 2.**
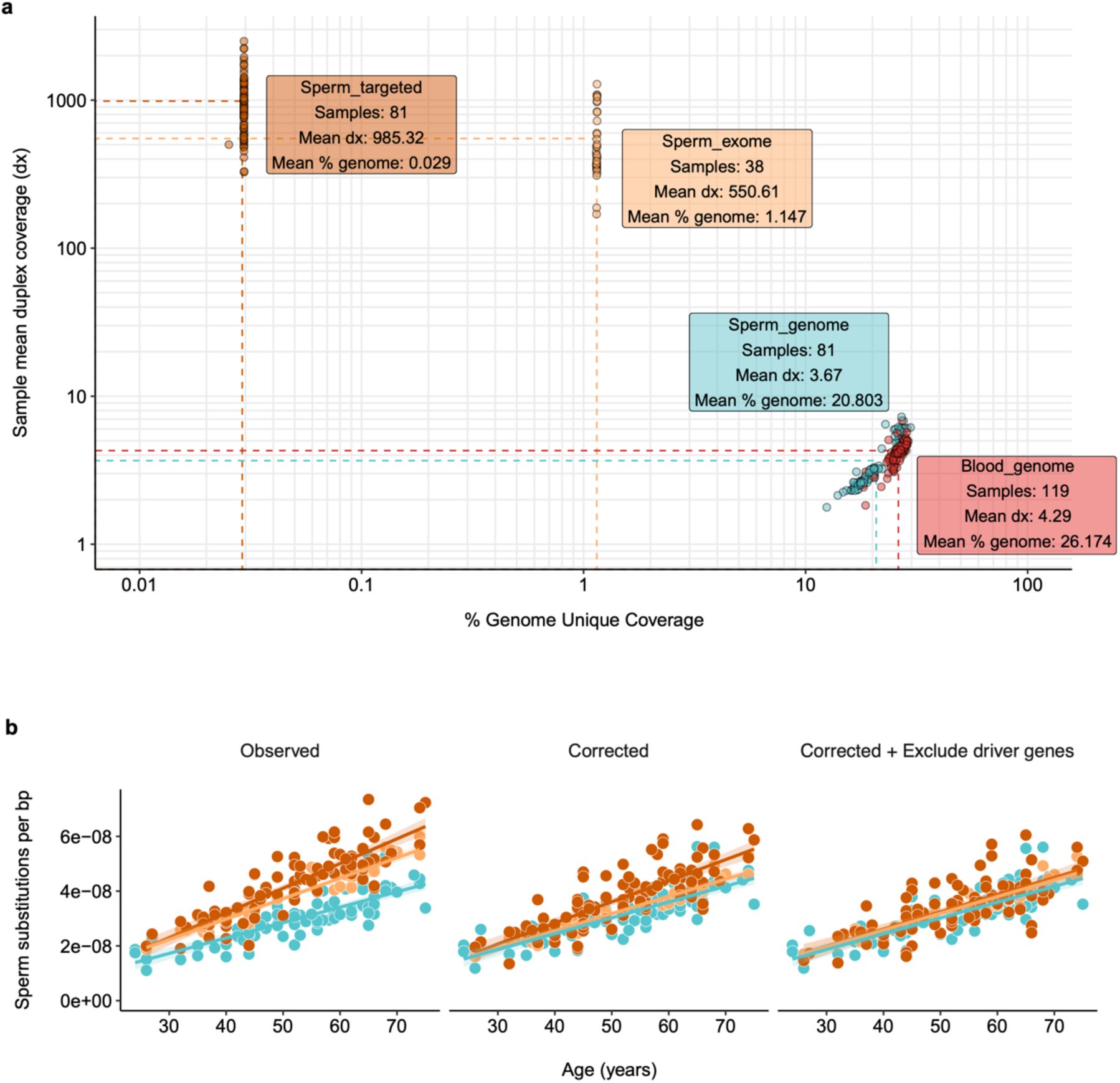
Coverage summary. **a**, Mean duplex coverage (log scale) and percentage of genome covered (log scale) per sample. Panels summarise the mean duplex coverage (dx) and mean percentage of genome covered per NanoSeq type and tissue. **b,** Mutation burden of targeted (dark orange), exome (yellow), and genome (blue) sperm sequenced samples that are observed without correction (left), corrected for trinucleotide composition of covered base pairs relative to the whole genome (middle) or corrected and masked for mutations and coverage in the 44 genes linked to germline positive selection (right). Model fits are linear regressions with 95% CI bands.

**Extended Data Fig. 3.**
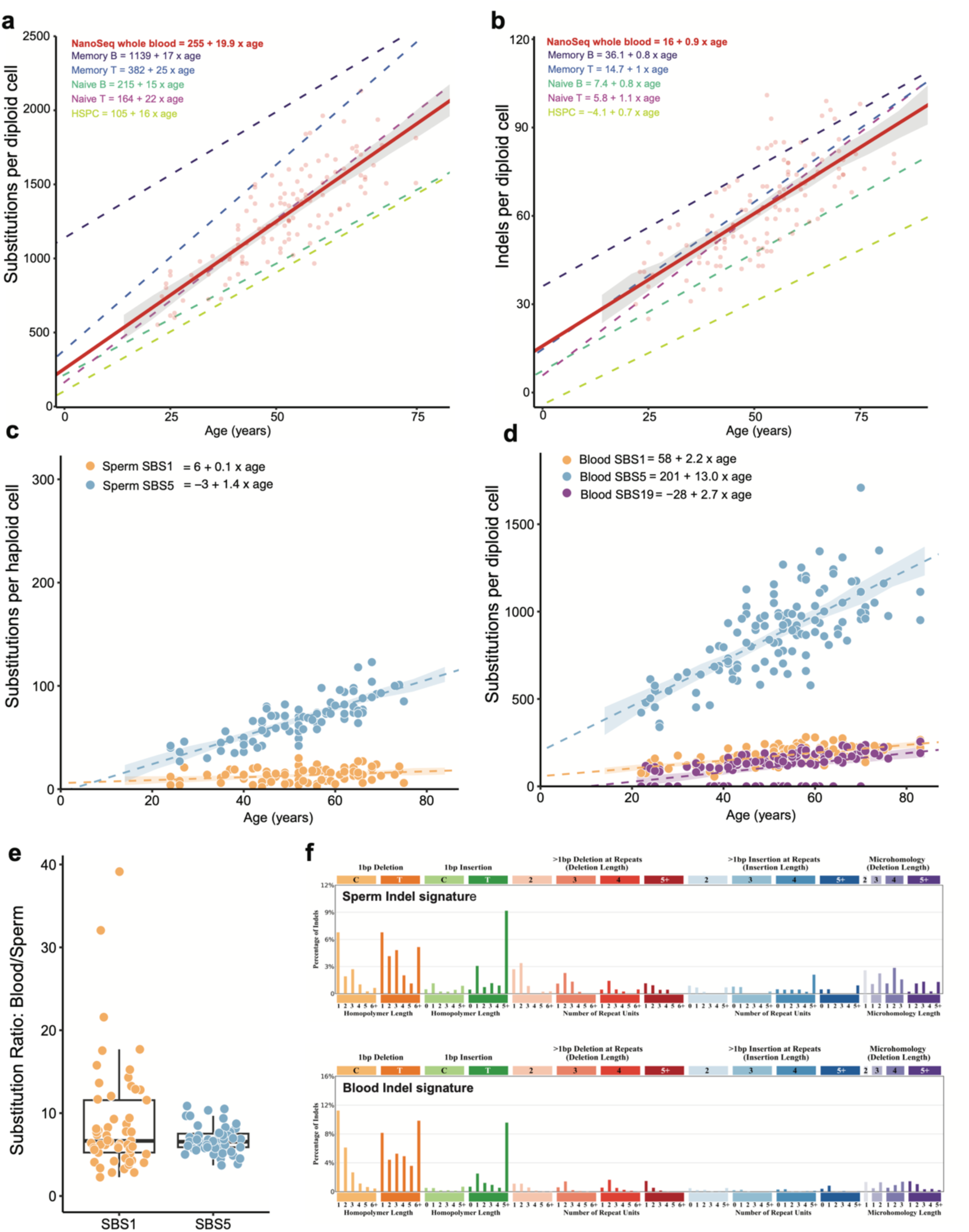
Mutation rates relative to blood cell types and split by signatures. **a,b,** Substitutions (**a**) and indels (**b**) per diploid cell from blood NanoSeq relative to specific blood cell types^8^**. c,d,** Substitutions per haploid cell for sperm (**c**) and diploid cell for blood (**d**) split by signature contributions of SBS1, SBS5, and SBS19. **a,b,c,d,** Models are linear mixed regressions with 95% CIs calculated by parametric bootstrapping. **e,** Ratio of age-corrected blood to sperm substitutions per diploid cell per year for mutations assigned to SBS1 and SBS5. Each dot corresponds to an individual with both a blood and sperm sample and where individuals had multiple timepoints the mean value of all timepoints in that tissue was used. Box plots show the interquartile range, median, and 95% confidence interval for the median. **f,** Distribution of indel types observed in sperm and blood.

**Extended Data Fig. 4.**
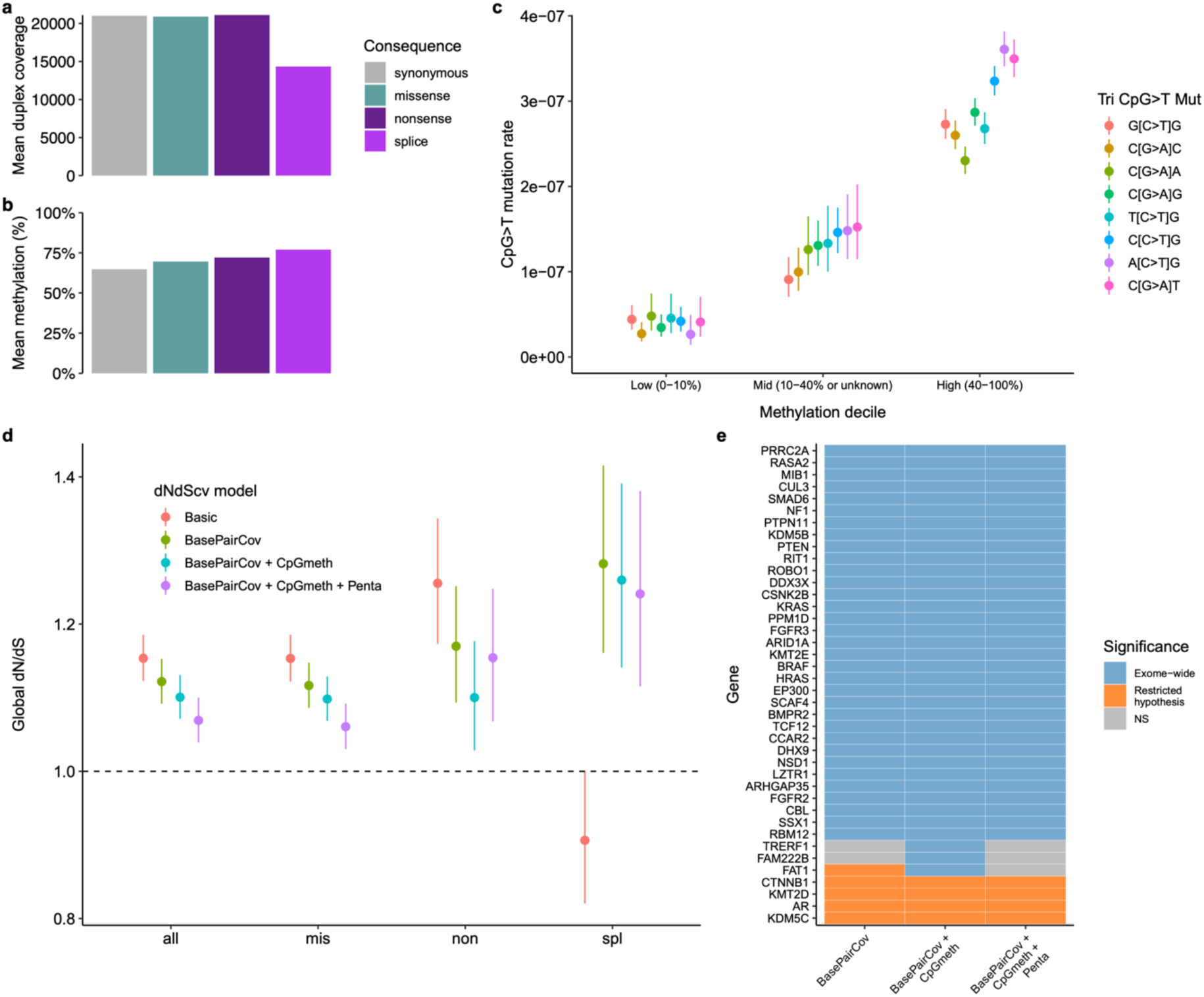
Model selection dN/dS. **a,b,** Mean duplex coverage (**a**) and methylation percentage (**b**) of all base pairs with exome sequencing coverage split by mutation consequence. **c,** C>T mutation rate at CpG sites in exome sequenced samples split by methylation bin based on percentage methylated from testis bisulfite sequencing^70^. **d,** Comparison of global dN/dS values from exome sequenced samples using different modifications to the *dNdScv* algorithm. Categories are all nonsynonymous mutations, missense, nonsense or essential splice. The basic model excludes genes which have no coverage but otherwise uses default parameters. Additional models show the impact of adding corrections for duplex coverage per base pair (BasePairCov), CpG methylation level (CpGmeth), and pentanucleotide context (Penta). **e,** Comparison of per-gene significance in exome-wide (blue) or restricted hypothesis (orange) dN/dS tests using the different models. Genes that did not reach significance in either test are shown in grey. Error bars indicate 95% CIs.

**Extended Data Fig. 5.**
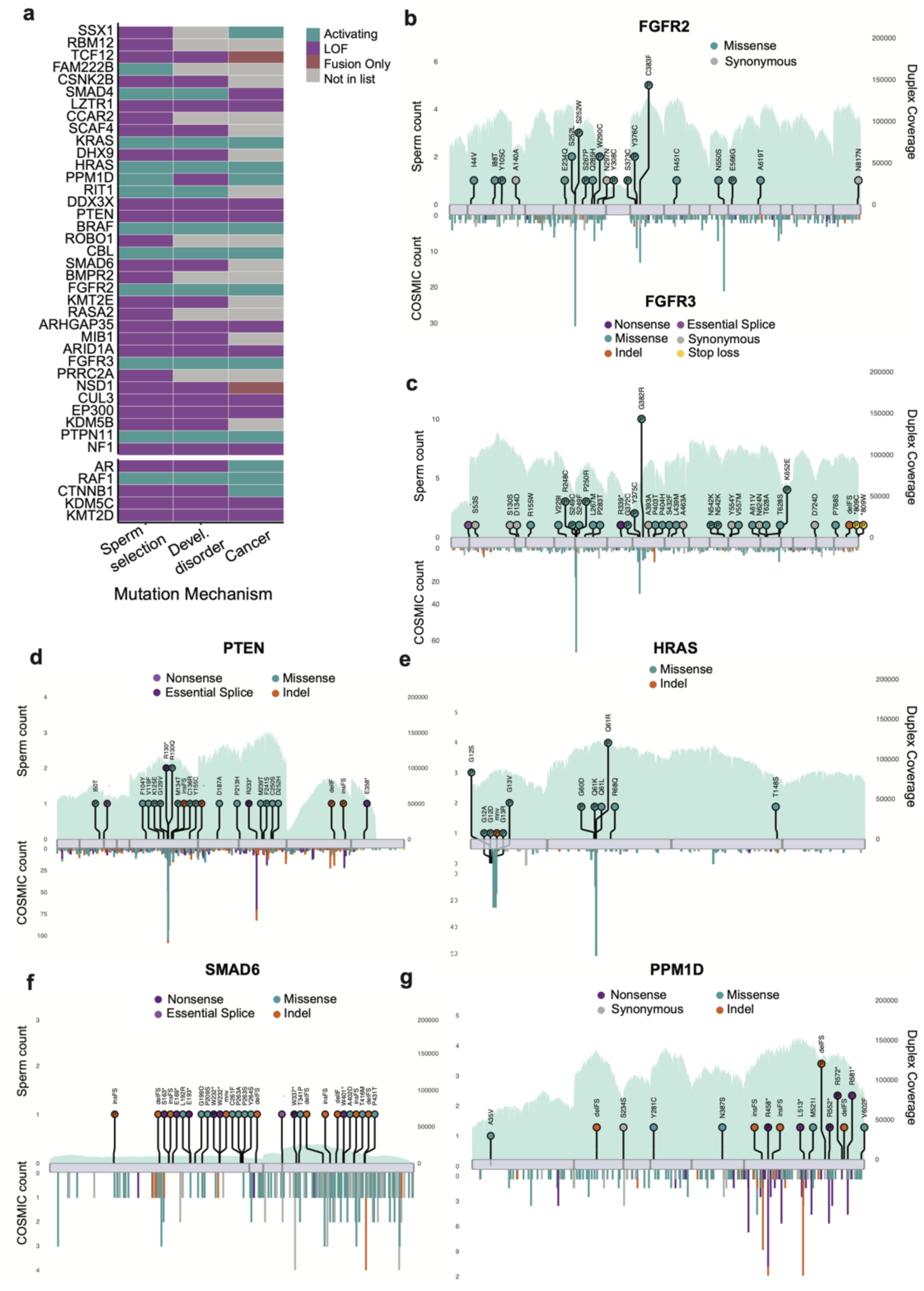
Gene mutation mechanisms. **a,** The mutation mechanism assigned to each gene based on the mutation pattern in sperm, developmental disorders and cancer (**Methods**). **b,c,d,e,f,g,** Observed sperm mutations across the cohort for six illustrative genes where the height of the “lollipop” represents the number of unique samples with a mutation at that location and the colour represents its mutation type. Mutations are labelled with their amino acid consequence for point substitutions or their insertion (ins)/deletion (del) consequence of in frame (IF) or frameshift (FS). A “P” indicates that the variant is classified as pathogenic/likely pathogenic in ClinVar^34^. Exons are shown as purple rectangles and the blue background represents the total duplex coverage across the cohort. Lines below the gene indicate COSMIC somatic mutations in cancer within that gene^32^.

**Extended Data Fig. 6.**
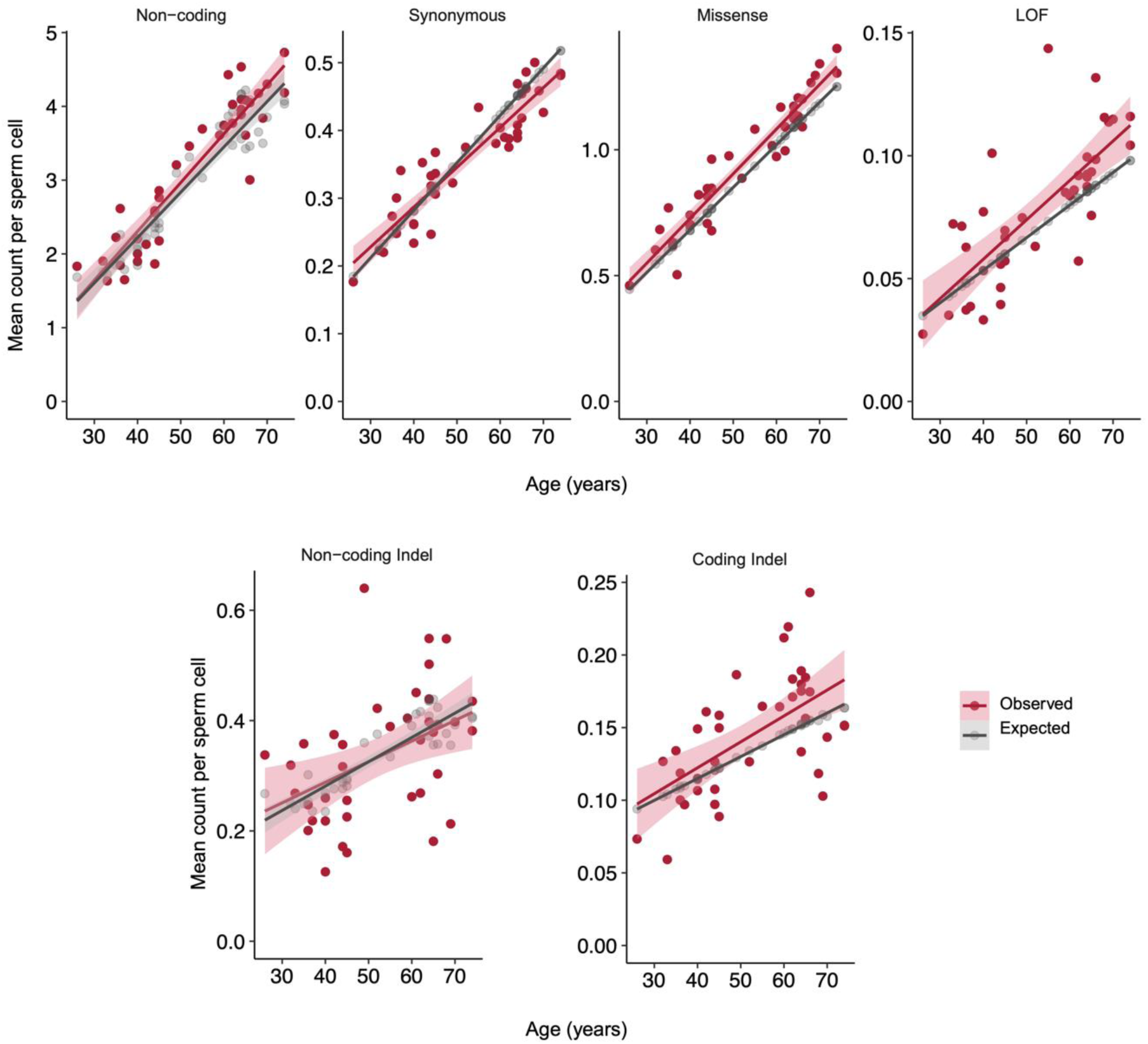
Mean variant class count per individual by age. The relationship between age and the mean count of SNVs (non-coding, synonymous, missense, and loss-of-function (nonsense or essential splice)) and indels (non-coding indel and coding indel) per sperm cell. The red points represent the observed values for each individual. The grey line represents the expected mutation count per sperm based on the germline mutation rate model. Error bands indicate 95% CIs of linear regressions.

**Extended Data Fig. 7.**
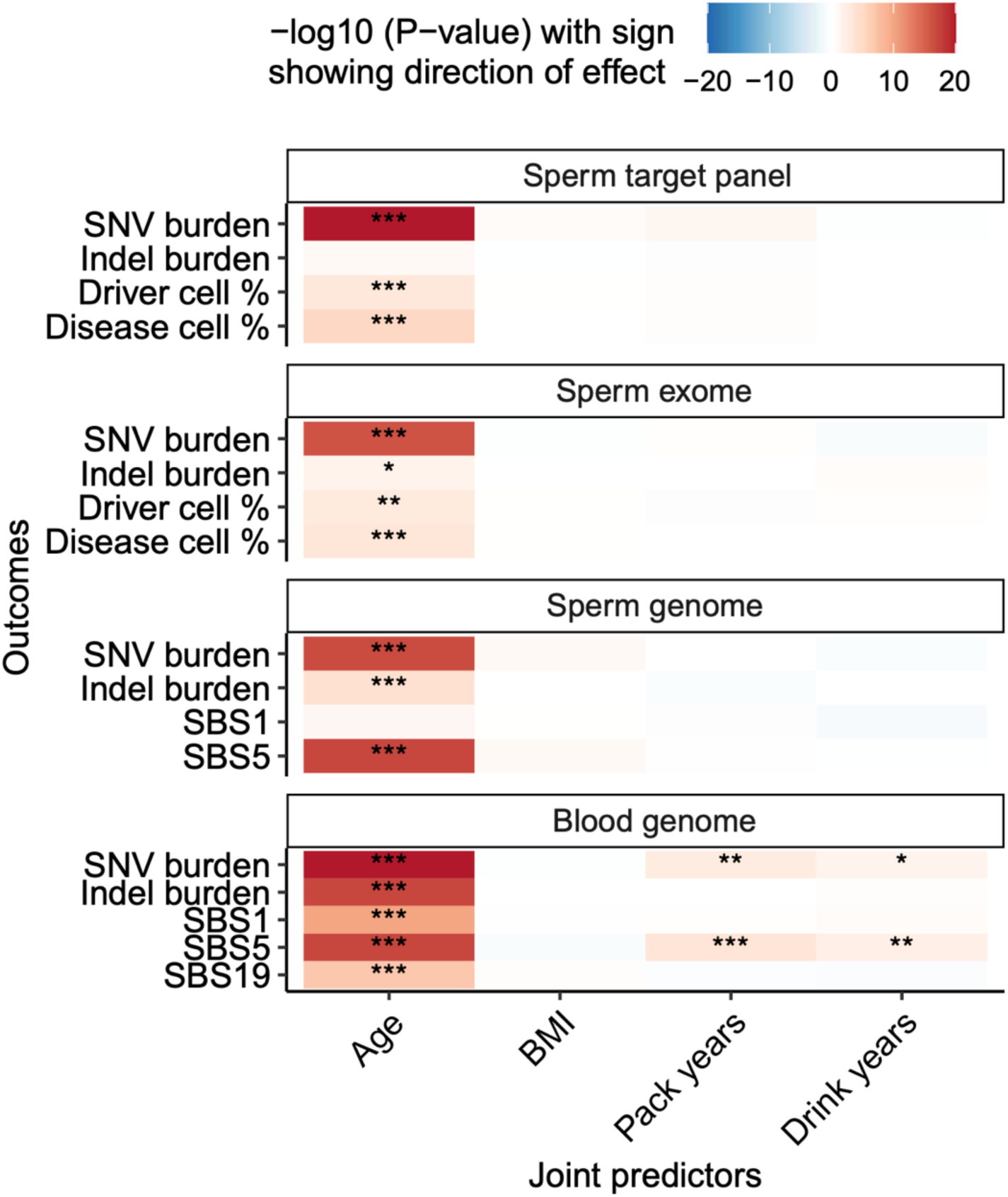
Phenotype correlations. Correlation of cohort phenotypes to mutation outcome variables, with different sequencing datasets split by facets. Joint predictor glm models used the gaussian family with FDR corrected *P* values. Asterisks indicate significance level of corrected *P* value: (\**P* value >0.01 to <0.05, \*\**P* value >0.001 to <0.01, \*\*\**P* value <0.001).

**Extended Data Table 1.**
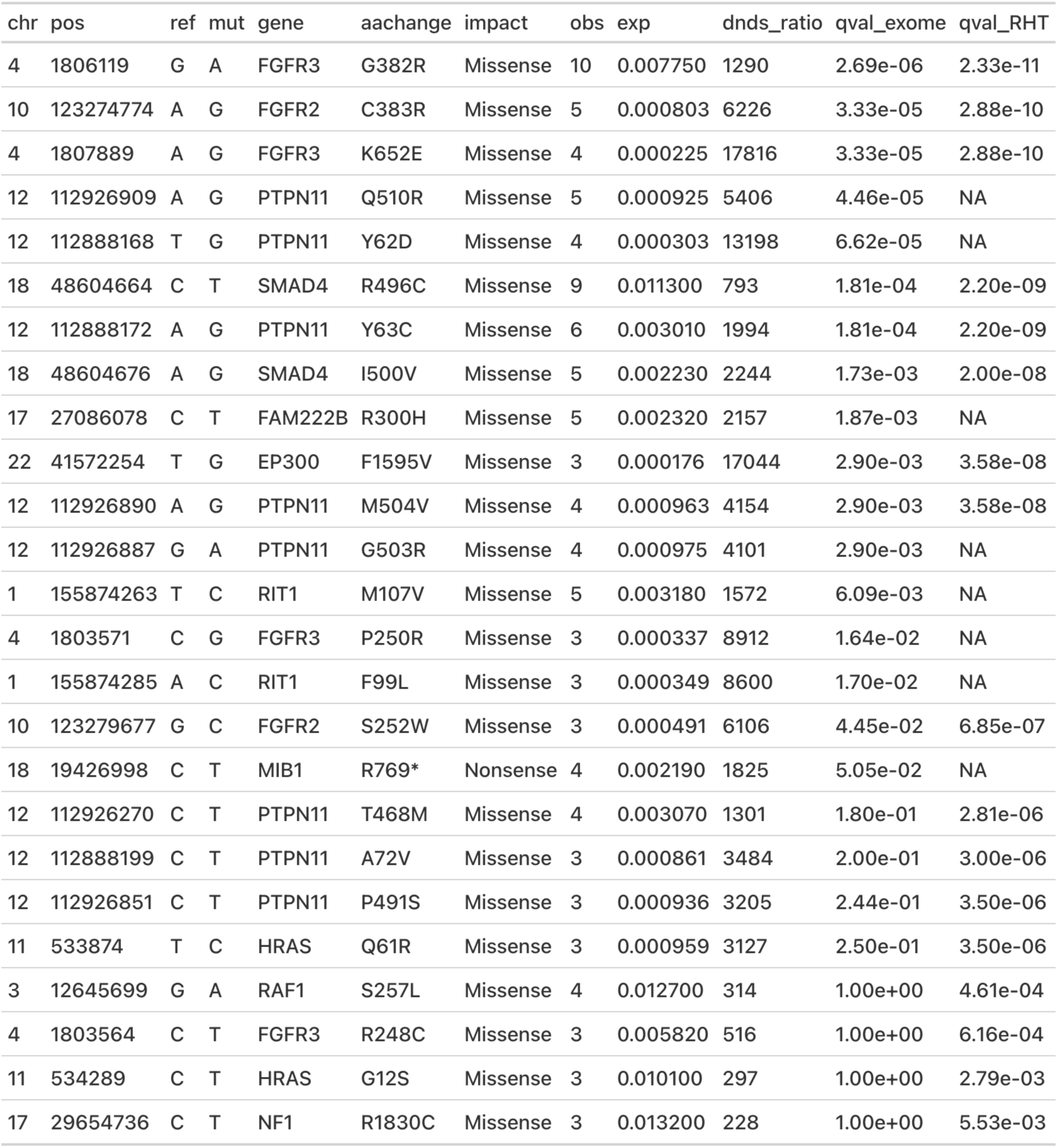
Significant SNV hotspots from dN/dS exome-wide and restricted hypothesis tests (RHT)

