## Supplementary Notes for "Sperm sequencing reveals extensive positive selection in the male germline"

### Supplementary Material for
### Sperm sequencing reveals extensive positive selection in the male germline

Matthew DC Neville[1], Andrew RJ Lawson[1], Rashesh Sanghvi[1], Federico Abascal[1], My H Pham[1], Alex Cagan[1], Pantelis A Nicola[1], Tetyana Bayzetinova[1], Adrian Baez-Ortega[1], Kirsty Roberts[1], Stefanie V. Lensing[1,2], Sara Widaa[2], Raul E Alcantara[1,3], María Paz García[4], Sam Wadge[4], Michael R Stratton[1], Peter J Campbell[1], Kerrin Small[4], Iñigo Martincorena[1], Matthew E Hurles[1], Raheleh Rahbari[1]

[1]Cancer, Ageing and Somatic Mutation, Wellcome Sanger Institute, Hinxton, United Kingdom
[2]Sequencing Operations, Wellcome Sanger Institute, Hinxton, United Kingdom
[3]Quotient Therapeutics Limited, Saffron Walden, UK
[4]Kings College London, Department of Twin Research & Genetic Epidemiology, London, United Kingdom

#### Table of Contents:

**Supplementary Note 1. Manual sperm counting**
**Supplementary Note 2. Mutation burden twins, timepoints, and intercepts**
**Supplementary Note 3. Running dN/dS and model selection**
**Supplementary Note 4. Potential influences on birth rate of positively selected variants**

#### Supplementary Note 1. Manual sperm counting

The single-molecule accuracy of duplex sequencing methods like NanoSeq renders mutation calls sensitive to non-target cell-type contamination[1]. A typical semen sample is composed predominantly of sperm, however it can also contain non-sperm cells such as immature germ cells, epithelial cells or leukocytes[2]. We performed cell counting to determine the concentration of sperm and non-sperm cells present in bulk semen samples sequenced, measured in millions of cells per ml (M/ml).

Manual assessment of semen cell concentration was performed using the Improved Neubauer Haemocytometer, as recommended by the World Health Organisation (WHO)[2]. Each frozen semen sample was liquified and mixed, and then 10µl of semen was visualised under ×200 magnification to approximate the number of sperm in the field of view. This approximation was used to determine the appropriate dilutions as described in Table 2.1 of the WHO manual[2]. Dilutions were then created with 50µl of semen and the previously determined dilution volumes of sterile water. The dilutions were then vortexed and 10µl was loaded into the Improved Neubauer Haemocytometer counting chamber. Counting from the haemocytometer was performed at ×200 magnification using the 25 large squares of the central grid.

A small number of samples were stained to visualise the difference in sperm concentrations using a Papanicolaou staining protocol[3] (**Extended Data Fig. 1a,b,c**). Briefly the semen was smeared onto a slide to create a thin layer of cells and then fixed. The slide was then submersed successively in the following reagents for the indicated time period: ethanol 50% for 30 seconds; ultra-pure water for 1 minute; Gill's Haematoxylin for 15 seconds; tap water wash for 15 seconds; tap water wash for 20 seconds; tap water wash for 20 seconds; ethanol 70% for 20 seconds; ethanol 100% for 40 seconds; ethanol 100% for 2 minutes; Orange G for 20 seconds; ethanol 100% for 2 minutes; Papanicolaou EA50 for 20 seconds; ethanol 100% for 20 seconds; ethanol 100% for 20 seconds; xylene for 20 seconds; xylene for 20 seconds; xylene for 20 seconds.

For sperm, counting was performed by row (5 squares), stopping at the end of a row if the count was over 150 sperm or if all rows (25 squares) had been counted. Round cells and epithelial cells were counted together and all 25 squares were always counted, however multiple epithelial cells were only found in a single azoospermic sample and therefore the concentration is primarily representative of round cells. Concentration of sperm and non-sperm cells in the sample was then calculated using the formula below where C is the concentration (Million/ml), N is the total number of sperm/non-sperm cells counted, n is the total number of rows observed and d is the dilution applied:

$$C = (N/n) \times (1/20) \times d$$

Of sperm samples counted, 5 were azoospermic (zero sperm present), 14 were oligozoospermic (<15M/ml) and 66 were in a normozoospermic range (>=15M/ml) (**Extended Data Fig. 1d**). The mean concentration of all samples was 75.7M/ml and the concentration was not significantly correlated to age ($p = 0.18$, Pearson's correlation). Cell counting of non-sperm cells present found all but 6 samples were below a concentration of 3M/ml and a mean concentration of 1.07M/ml. Other than azoospermic samples we found 6 samples with a ratio of sperm to round cell concentration below 10 to 1.

The mutation burden for each sample sequenced was calculated by dividing the number of mutations called by the total duplex coverage of the sample. After dividing by the age at which the sample was collected to account for the linear correlation of mutation rate with age, most samples showed mutation burdens per year (substitutions/bp/year) consistent with their tissue type (**Extended Data Fig. 1e**). The five azoospermic samples however, had outlier mutation burdens which were comparable to the blood burden per year (**Extended Data Fig. 1e**).

Examining non-azoospermic samples we find that only one of the samples with a ratio of sperm to round cells below 10 is an outlier in the relationship between mutation burden with age. This outlier sample was the sperm sample with the lowest non-zero sperm concentration (0.35M/ml) and the lowest non-zero sperm to round cell ratio (1.75), consistent with low level somatic cell contamination. Based on these findings we excluded the five azoospermic samples and the outlier oligozoospermic sample from subsequent analyses.

The mutation burden estimate in sperm presented in **Figure 1** of the main text was 1.67 haploid single base substitution (SBS) per year (95% confidence interval (CI) = 1.41-1.92, linear mixed-effect regression) with an intercept of 3.0 haploid SBS (95% CI = -10.3 to 16.4). This estimate is not not substantially impacted by excluding samples with oligozoospermia: 1.68 haploid SBS per year (95% CI = 1.38-1.98, linear mixed-effect regression) and intercept of 3.1 haploid SBS (95% CI = -12.2 to 18.5) or by excluding those samples with a ratio of sperm to non-sperm cells below ten: 1.68 haploid SBS per year (95% CI = 1.41-1.94, poisson regression) and intercept of 3.4 haploid SBS (95% CI = -10.2 to 17.1).

#### Supplementary Note 2. Mutation burden twins, timepoints, and Y-intercepts

Within the cohort, there were a total of 8 monozygotic (MZ) twin pairs and 3 dizygotic (DZ) twin pairs passing sample quality control. The cohort also included individuals with 1-3 timepoints in blood and 1-2 timepoints in sperm (**Supplementary Table 1**). In 51 individuals with two blood timepoints we found that for 33 individuals (65%) the mutation burden increased between the first to second timepoint and that all 6 individuals with a third blood timepoint had an increased burden from timepoint two to three. From 24 individuals with two sperm sample timepoints, all but one had increased mutation burden in the second timepoint. The higher variability between timepoints in blood is likely a reflection of a smaller average time gap between sample collection (average of 12.1 years in sperm vs 8.1 years in blood) and possible heterogeneity in cell composition sequenced in different blood samples and across the timepoints.

We examined the ΔAIC of simple linear mutation burden vs age models to models that account for timepoints or twin status using random effects to test which better models the data. Specifically the models and equation used were ΔAIC = AIC(lm(burden ~ age)) - AIC(lmer(burden ~ age + (age - 1|ID), REML = FALSE)), where burden was for either sperm or blood and ID was for either individuals or twin pairs. We found that both random effects models produced a worse fit for sperm with ΔAIC of -2.0 and -0.1 for twins and timepoints respectively. In contrast we found that both models provided a significantly better fit in blood with ΔAIC of 10.1 and 7.1 for twins and timepoints respectively. The lack of predicted effect in sperm could be partly due to the lower mutation burden in sperm compared to blood, reducing statistical power.

We also note that the observed lower Y-intercept of mutation burdens in seminiferous tubules compared to sperm is likely due to the technical limitations and biological factors. In the seminiferous tubules, the Laser Capture Microdissection (LCM) method identifies mutations that are clonal, which explains the negative Y-intercept observed for the testes data. Previous studies estimate that it takes approximately 15.4 years (95% CI: 5.7 to 23.7) for the seminiferous tubule populations to become clonal[32]. This is because the LCM approach primarily detects mutations in the spermatogonial stem cells (SSCs), which are the self-renewing cells that give rise to all sperm. The clonal mutations identified in this context reflect mutations that occurred early in life and have expanded in the SSC population over time.

In contrast, mutations observed in the sperm data and in trio DNMs are impacted not only by mutations in SSCs but also by additional mutations that arise during the differentiation process of spermatogenesis. The NanoSeq method used in the sperm analysis can detect these mutations, including those that occur in the later stages of spermatogenesis and in mature spermatozoa, which would not be captured by the LCM-based study of testes. As a result, the sperm data includes both clonal mutations from SSCs and those accumulated during the development and maturation of sperm, leading to a higher Y-intercept in the sperm cohort.

#### Supplementary Note 3. Running dN/dS and model selection

Input mutations

When running all dN/dS calculations, to conservatively avoid counting possible single mutational events more than once, mutations shared within an individual's multiple timepoints were collapsed to single entries. Where estimates were for global dN/dS or gene set dN/dS only exome sequenced samples were used to avoid coverage bias. For tests examining the significance of individual genes, both the exome and targeted sequencing data were used as the differential coverage is accounted for in the duplex coverage adjustment.

Model selection

When calculating dN/dS it is important to account for systematic biases in mutation rate, particularly any biases which differentially impact synonymous vs nonsynonymous mutations. For this reason the *dNdScv* algorithm corrects for 192 mutation rate parameters, which represent the 96 trinucleotides and the transcribed vs non-transcribed strands[4]. This controls for the known differences in mutation rates between trinucleotides, which differ in their frequency for synonymous and nonsynonymous mutations due to codon biases. To ensure that the dN/dS values we calculated are not substantially impacted by any other such biases we investigated 3 possible sources of bias: sequencing coverage, methylation level at CpG sites, and extending mutation context correction from trinucleotides to pentanucleotides.

Each of these investigations involved modification of the mutation opportunities matrices that are central to the *dNdScv* algorithm. Specifically, each gene has a mutation opportunities matrix which by default has dimensions of 192x4 where the 192 rows represents the possible mutation contexts and the 4 columns represent the 4 substitution consequences considered: synonymous, missense, nonsense, and essential splice. The entries to the matrix are the number of opportunities for that mutation type at that consequence. For instance the [1,1] entry of each of these matrixes is the number of possible AAA>ACA mutations that cause a synonymous variant in that gene, while the [100,3] entry will be the number of GAC>GCC mutations that cause a nonsense mutation in that gene. These matrices are either combined across genes to calculate global dN/dS values or used on the gene level to compare the observed mutations in genes to the expected count based on the number of possible mutation opportunities.

*Coverage correction*

We first investigated the impact of correcting for differential coverage at the base pair level. The single molecule accuracy of duplex sequencing provides the exact number of DNA molecules where a mutation was callable, which we leveraged for this modification. To achieve this we implemented a custom per base pair coverage correction where, rather than using an opportunity matrix with the number of each trinucleotide per gene, we multiplied each matrix entry by its exact coverage in the cohort. For instance, if there were 4 possible AAA>ACA mutations in a gene that could cause a synonymous mutation, rather than multiplying having a 4 in the matrix at this position we added up the exact coverage at those 4 possible sites (e.g. 25,000 + 26,000 + 18,000 + 0). In this example the 4th site had no coverage either because that base pair was not sequenced or that site was masked in variant calling. Using this modification therefore has three advantages: (1) masking sites where a mutation was not callable, (2) correcting for differential coverage between genes, and (3) correcting for differential coverage within genes.

There were a number of germline polymorphism sites and known artefactual sites that were masked during variant calling (**Methods**), which, due to being skewed towards synonymous sites which are more common in population polymorphisms, can falsely inflate dN/dS values[4]. This could explain why, after implementing this adjustment to *dNdScv*, we observed reduced global dN/dS ratios for missense, nonsense, and all non-synonymous mutations overall (**Extended Data Fig. 4d)**.

Interestingly, we observed that essential splice sites, unlike other mutation types, had the global dN/dS ratio change from below 1, to levels comparable to other studies[5]. Upon further investigation, we found that, on average, splice variants had less coverage than other variant classes at a mean coverage of 14,340 compared to synonymous, missense and nonsense variants at 21,008, 20,900, and 21,108 respectively (**Extended Data Fig. 4a)**. This means that splice sites would have less chances to have a mutation call and therefore have a downwards biassed dN/dS value without this correction.

*Methylation level*

Next, we examined the possible influence of methylation level as a known strong influence on mutation rate at CpG sites. Comparing the mutation rate to methylation level assessed in testis[6], where the majority of germline mutations are thought to occur, we found a strong correlation between methylation percentage and mutation rate (**Extended Data Fig. 4c**). In theory this might not be an issue if the different mutation classes shared a similar distribution of methylation levels, however this was not the case. We find that across the covered regions of the exome, CpG sites where a synonymous variant was possible had the lowest methylation level (64.8%), followed by missense (69.6%), nonsense (72.1%), and essential splice sites (77.0%) (**Extended Data Fig. 4b**). If methylation level increases mutation rate and synonymous sites have a lower average methylation level then this will bias dN/dS estimates upwards. To correct for this we extended the mutational opportunities matrix from 192x4 to 208x4, where each of the 8 possible CpG trinucleotide opportunities was split into 3 rows based on percent methylation at that site, one each for low (0-10% methylation), mid (11-40% OR unknown), and high (>40% methylation) methylation. This correction slightly lowered the observed dN/dS ratios as expected (**Extended Data Fig. 4d**).

*Pentanuclotide context*

It has been shown that trinucleotide models accurately capture most context-dependent mutational biases in dN/dS models for cancers, with the exception of both melanoma and *POLE* hypermutator tumours where known pentanucleotide context dependence biassed trinucleotide models[4]. Several papers have shown through population data resources that extended contexts beyond trinucleotides (3mer) such as pentanucleotide (5mer) and heptanucleotide (7mer) contexts impact germline mutation rates[7,8]. The size of the dataset did not allow investigation of heptanucleotide context, but we extended the *dNdScv* model from trinucleotide to pentanucleotide context to evaluate its impact. This was done by extending the trinucleotide + methylation opportunity matrix from 208x4 to a pentanucleotide + methylation matrix of 3328x4. This is a 16 fold increase in contexts from each of the four possible nucleotides up and downstream.

We found that this change resulted in lower estimated global dN/dS estimates driven primarily by lower missense enrichment (**Extended Data Fig. 4d**). To investigate which extended contexts had mutation rate changes at extended contexts we calculated the difference in rates between trinucleotide and pentanucleotide contexts using the confidence intervals to estimate the standard errors (**Supplementary**

**Table 9)**. A two-tailed z-test was performed for each comparison, and the resulting p-values were adjusted for multiple testing using the Benjamini-Hochberg procedure to control the false discovery rate. We found 51 significant changes in pentanucleotide context relative to trinucleotide context, with a large portion of those (22/51) being variability within extended contexts of A[T>C]T (or A[A>G]T) as previously identified[7,8].

###### *Impact on gene level tests*

All three adjustments to the model had notable impacts on global dN/dS values, but generally had only minor impacts on gene level significance. For instance, there was very little variability in which genes reached significance exome-wide or with the restricted hypothesis test in each of the three model adjustments (**Extended Data Fig. 4e**). Specifically, the genes significant under the base pair coverage model were almost identical to those in the final chosen model of base pair coverage + CpG methylation + pentanucleotide, with only the gene *FAT1* being discordant in the restricted hypothesis test. The second model with base pair coverage + CpG methylation had a few differences in significant genes, with *FAT1*, *TRERF1*, and *FAM222B* reaching exome-wide gene significance, however *FAM222B* is significant in the hotspot tests and therefore was retained on the significant gene list even in the chosen base pair coverage + CpG methylation + pentanucleotide model. The finding that the global dN/dS model changes are more impacted by model adjustments is consistent with the understanding that small adjustments in mutation rates have a larger impact when propagated across the exome than over a smaller region like an individual gene.

##### Hotspot tests with sitedNdS

As some driver genes are mutated at specific hotspots rather than across the entire gene, we additionally applied the *dNdScv* sitedNdS function[9] that detects positive selection at specific base pair sites. This algorithm was modified to account for base pair level duplex coverage similarly to the gene level test by using a dN/dS model fit with base pair level correction and adjusting for the relative coverage of each recurrently observed mutation tested. The sitedN/dS test was conducted exome-wide and on a set of 1,236 known hotspots from TCGA[10] and the MSK pan-cancer cohort[11] and 732 sites with 2 or more variants in the Deciphering Developmental Disorders (DDD) cohort[12] not already in the cancer hotspot list (32 overlapped). Sites and their genes were considered under positive selection if a site reached an FDR q-value $< 0.1$.

##### Driver mutation estimates

The number of driver mutations in the exome dataset explained by known driver genes was estimated by comparing the number of excess non-synonymous substitutions assigned by *dNdScv* across the exome to those found in genes linked to germline positive selection. For each test we calculated the estimated number of drivers (nD) = $((\omega NS-1)/\omega NS)nNS$, where $\omega NS$ is the estimated dN/dS ratio of non-synonymous mutations and nNS is the observed number of non-synonymous mutations. The fraction of explained drivers was then given by $nD_{selection}/nD_{exome}$ with confidence intervals calculated through propagated uncertainty. The reported estimate was given using the custom *dNdScv* model with duplex base pair, CpG methylation, and pentanucleotide corrections, and thus the estimate, due to changes in $\omega NS_{exome}$ (**Extended Data Fig. 4d**) will be sensitive to the model used.

<u>External germline dataset dN/dS</u>

We calculated estimated global dN/dS values for external germline mutation datasets using the basic *dNdScv* model and the genes with coverage in sperm exomes. Control DNMs were downloaded from denovo-db v.1.6.1[13]. Developmental disorder DNMs were from the 31k developmental disorder trio cohort[12]. Population variants were downloaded from gnomAD v2.1.1 exomes[14] and split by reported allele frequencies.

<u>Gene set enrichment</u>

Gene set enrichment was conducted in the same way as global dN/dS calculations using *dNdScv* and the basePairCov + CpG meth + Penta custom model, with the relevant set of genes input as the 'gene_list' parameter. Expression gene sets examined were expression levels from 7 bins of mean expression levels across germ cell stages and expression clusters of genes most characteristic to certain germ cell stages[15]. Pathway gene sets examined were the 44 germline selection genes found here or previous and cancer gene census genes[16] split by ten canonical cancer pathways in KEGG[17].

<u>Mutation set enrichment</u>

Mutation set enrichment of cancer and developmental disorder mutations was achieved by comparing observed counts of mutations to expected counts. Cancer somatic mutation counts were from the exome and genome wide screens of the COSMIC[16] (v99) and developmental disorder mutations were from the 31k developmental disorder trio cohort[12]. Expected counts were generated by multiplying the cohort wide coverage at each possible SNV matching the relevant database of mutations by the mutation rate specified in the custom basePairCov model developed from *dNdScv*.

#### Supplementary Note 4. Potential influences on birth rate of positively selected variants

It is important to recognise that not all positively selected or disease-causing mutations observed in sperm will necessarily appear at the same rates in live births. This note outlines some possible mechanisms and case examples where positively selected mutations may be reduced in frequency before reaching live births. However, future studies will be needed to prove and quantify these effects.

Although not key to interpreting sperm mutations in this study, future studies that investigate mutations in testis should consider that some driver mutations may favour clonal growth of spermatogonia without leading to full differentiation into functional sperm. Such mutations might provide a growth advantage by promoting proliferation while impairing or completely blocking spermatogenesis.. It has already been shown that many driver mutations in the testis have reduced spermatogenesis relative to adjacent wild-type tubules[18]. However, many of those mutations were observed to be under positive selection in this paper, indicating that this reduced spermatogenesis is not sufficient to fully counteract the clonal expansion. Further studies will be needed to determine whether any clonal expansion causing mutations in the testis exist for which severely reduced or completely impaired spermatogenesis are sufficient to counteract positive selection in the testis. Additionally, with the many new positively selected genes uncovered in this work, future studies could investigate the degree to which reduced spermatogenesis does or does not influence the frequency of different driver mutations observed in sperm.

For positively selected variants which are compatible with spermatogenesis, such as those identified in sperm from this study, the next stage at which they may reduce sperm fitness is the ability to successfully fertilise an egg. It is unclear whether mutations which are compatible with spermatogenesis would then have a frequent impact on the ability of sperm to fertilise an egg, given that the pathways implicated in fertility are generally different than those which confer growth advantages to cells. For instance, of the 104 genes with moderate, strong, or definitive links to male fertility[19], only *AR* is also a significantly positively selected gene in sperm. Even in the case of *AR*, its link to fertility is pre-spermatogenesis via reproductive organ development or meiotic arrest through sertoli cells.

In contrast, it is intuitive that mutations which alter cellular differentiation in the testis might be disruptive to the tightly regulated developmental cell divisions of embryogenesis. Those that are disruptive enough may even lead to pregnancy loss, which would result in a lower number or none of certain positively selected mutations appearing in live births. It is known that 8–15% of clinically recognised pregnancies and approximately 30% of all pregnancies are lost prior to birth[20–22]. Although the majority of miscarriages with a genetic cause are due to chromosome abnormalities[23], non-aneuploid genetic causes from germline mutations are also observed[24]. The contribution of non-aneuploid DNMs to pregnancy loss has not been systematically quantified, but there are several lines of evidence that suggest paternal DNMs are involved in some fraction of cases. These include the finding that advanced paternal age, the main risk factor for DNMs and positively selected variants in sperm seen in this work, is a 2.05 (95% CI, 1.06–2.20) risk factor for miscarriage independently of maternal age[25]. Additionally, there are case reports of positively selected sperm variants among the likely causal variants in exome sequencing studies of pregnancy loss, including the Noonan syndrome associated *PTPN11* A72T mutation[26] seen in the sperm of two individuals in this work and the *FGFR3* R248C thanatophoric dysplasia mutation[27] seen in the sperm of 3 individuals. An intriguing set of mutations that we observed in this study are the G469V in *BRAF*, C383R in *FGFR2*, and A72V in *PTPN11* mutations that were all observed multiple times in sperm and are frequently observed in cancers but to our knowledge have never been observed in a live birth. These three variants potentially fit into a previously hypothesised pattern of mutations that confer a selective advantage that is strong enough that they are only observed in cancers and are not compatible with developing to live birth[28].

It is worth noting that as screening techniques for pregnancies have become more advanced and widely adopted, some positively selected sperm mutations are likely being selected against prior to birth through elective pregnancy terminations. For instance, the achondroplasia and thanatophoric dysplasia causing sites in *FGFR3* as well as multiple *PTPN11* and one *RIT1* Noonan syndrome pathogenic sites have been diagnostic variants in a prenatal sequencing study of foetuses with anomalies detected by ultrasonography[29]. Another study found causal DNMs in foetuses terminated after prenatal testing for variants in five of the sperm positively selected genes: *RIT1*, *ARID1A*, *RAF1*, *FGFR2*, *DDX3X* [30]. As prenatal sequencing becomes more precise and less invasive, such as in recent advances for prenatal screening from cell-free DNA[31], this may lead to more elective terminations in such cases.
